# Current practices, challenges, and future directions in subcutaneous oncology monoclonal antibody delivery: A qualitative study

**DOI:** 10.64898/2026.05.06.26352582

**Authors:** Chris Franzese, Madeleine Anderson, Jenny Wu, Abhi Raj, Marty Coyne, Sherri Biondi

## Abstract

**Background:** Subcutaneous oncology monoclonal antibodies (SCOmAbs) offer significant benefits compared to intravenous formulations, including reduced administration time and potential for self-administration. However, little has been published on real-world preparation and administration practices with these novel formulations.

**Methods:** This was a qualitative, exploratory study of 30 participants (10 patients receiving SCOmAbs, 10 nurses, and 10 pharmacists/pharmacy technicians) across various U.S. healthcare facilities. One-on-one, in-depth interviews examined current practices, pain points, device integration potential, projected impact of increased SCOmAb adoption, and perspectives on home administration.

**Results:** Findings revealed considerable variability in SCOmAb preparation and administration practices. While preparation methods generally aligned with typical parenteral workflows, notable deviations included bedside preparation by nurses (7/30), use of syringe pump modules (3/19), and one case of patient self-administration at home. Most participants utilized closed-system transfer devices (12/22) despite inconsistent hazardous drug treatment between facilities. Administration challenges included ergonomic difficulties for nurses during manual push (5/9 reporting physical discomfort) and variable injection techniques to accommodate patient comfort. Nurses reported significant workflow impact from being “tethered” to patients during administration, which could require staffing adjustments as SCOmAb frequency increases. Most patients (6/9) expressed interest in home administration for potential time savings and flexibility, though concerns about training, support, and safety were common.

**Conclusions:** As SCOmAb utilization expands, facilities may face barriers associated with increased demand that could necessitate innovative solutions. To leverage the full potential of SCOmAb products, developers should consider how next generation product presentations might minimize identified pain points, streamline administration, and facilitate safe transition to home self-administration. Delivery device integration, whether through prefilled syringes, portable infusion pumps, or other delivery systems, may address current challenges but requires careful consideration of facility infrastructure, product complexity and usability, workflow impact, and patient training requirements.

## Introduction

The transition from intravenous (IV) to subcutaneous (SC) delivery of biotherapeutics, such as monoclonal antibodies (mAbs), is prominent for a variety of reasons. Perhaps most impactful is the care setting flexibility, specifically self-administration at home, enabled by SC formulations that provides a host of patient benefits, including freedom, autonomy, and reduced time in clinic.[1,2] When at-home administration is not an option, SC delivery in clinic still offers significant advantages for healthcare providers (HCPs) and the broader healthcare system, including streamlined workflow, faster patient turnaround, and reduced overall treatment costs.[3–5] The benefits of SC administration may be most tangible among patients with cancer, as visits to the clinic for IV therapies are known to be associated with significant patient workload and treatment burden.[6] In this context, SC oncology mAbs (SCOmAbs), such as SC formulations of rituximab (Rituxan Hycela^®^, Genentech), trastuzumab (Herceptin Hylecta^®^, Genentech), daratumumab (Darzalex Faspro^®^, Janssen), the fixed-dose combination of pertuzumab and trastuzumab (Phesgo^®^, Genentech), atezolizumab (Tecentriq Hybreza^®^, Genentech), nivolumab (Opdivo Qvantig^®^, Bristol-Myers Squibb), pembrolizumab (Keytruda Qlex^™^, Merck) and amivantamab (Rybrevant Faspro^™^, Janssen; a bispecific antibody)[7] offer the potential for significant time savings for both patients and HCPs,[8–10] reduced systemic administration-related adverse effects,[11–14] and overall patient and HCP preference compared to their IV alternatives.[9,15–18]

Unlike most subcutaneously administered therapies, however, many existing SCOmAbs require larger injection volumes than what has been historically considered possible by conventional SC administration recommendations.[19] This is facilitated in part by co-formulation with recombinant hyaluronidase, which degrades hyaluronic acid in the extracellular matrix and allows for relatively rapid administration of large volumes into single SC injection sites.[20,21] Current standard of care for SCOmAbs is HCP administration in clinic by manual syringe injection (“manual push”) into the abdomen or thigh over several minutes.[22] While specific configurations and instructions vary for each product,[23–30] all SCOmAbs are currently supplied in standard vials, and HCPs are instructed to withdraw the full dose into a single syringe using a transfer needle, remove the transfer needle, and cap the syringe to await administration. Administering HCPs are advised to refrain from attaching a hypodermic injection needle or SC administration set until immediately prior to administration to avoid any clogging of the medication in the needle, and maximum in-syringe dwell times are noted for each product. In some, but not all cases, specific characteristics of recommended syringes, transfer needles, administration needles, and/or SC infusion sets are also specified. The dose volume(s), injection time(s), injection site(s), specified device characteristics, and in-syringe stabilities for on-market SCOmAbs as of the time of this manuscript preparation are summarized in Table 1. One additional area of handling variability may depend on whether or not these agents are considered hazardous drugs per facility protocols, as this decision has the potential to greatly influence both preparation and administration practices. Although no on-market SCOmAb currently appears on the National Institute for Occupational Safety and Health (NIOSH) List of Hazardous Drugs in Healthcare Settings (“NIOSH List”) in the US,[31] facilities may still use supplies like closed-system transfer devices (CSTDs) to protect healthcare personnel from drug exposure,[32,33] likely to preserve workflow consistency for all oncology products. However, using CSTDs to prepare and administer biotherapeutics has been the subject of significant controversy within the pharmaceutical industry, as doing so can potentially compromise biologic stability or full dose delivery.[34–36]

**Table 1.**
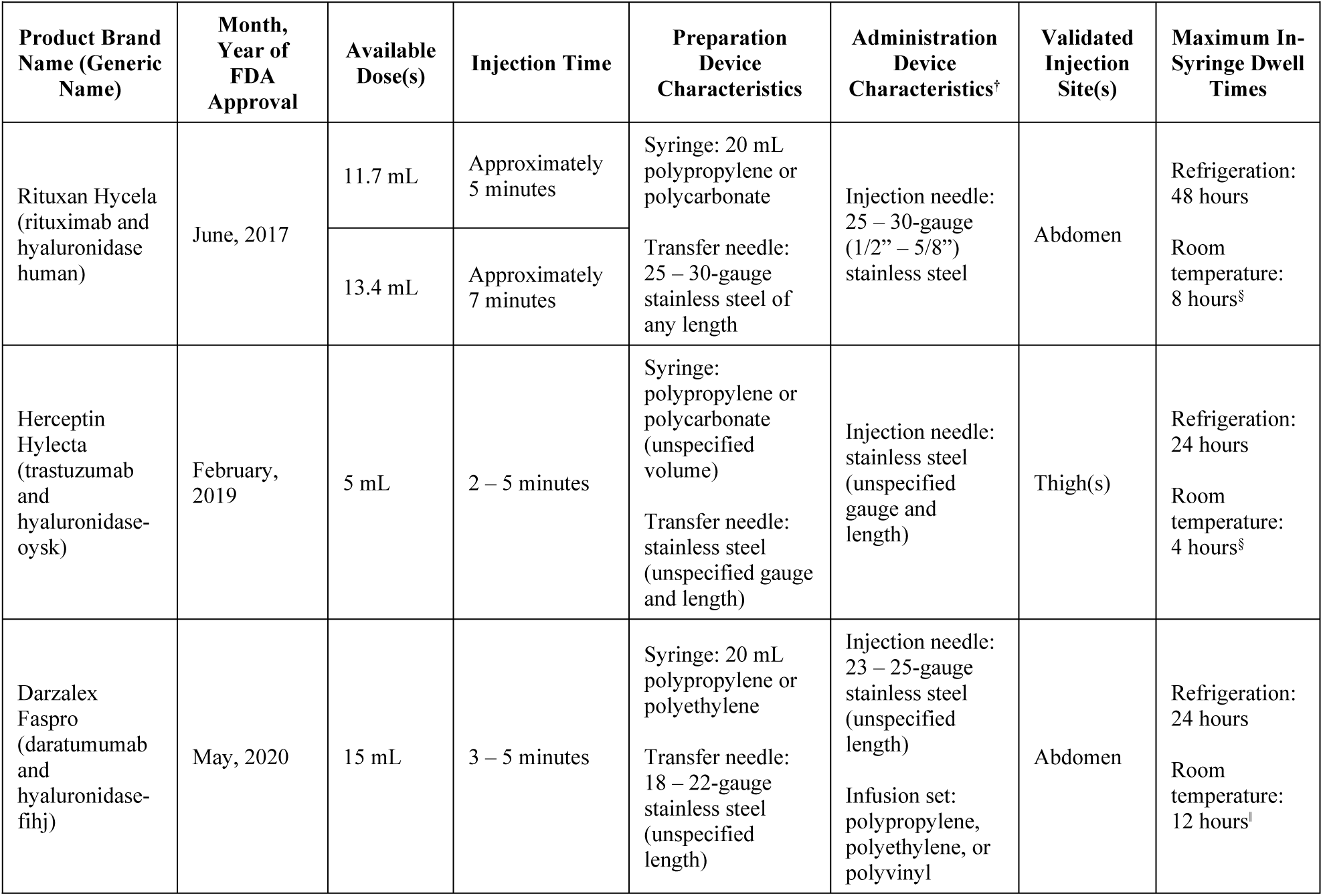
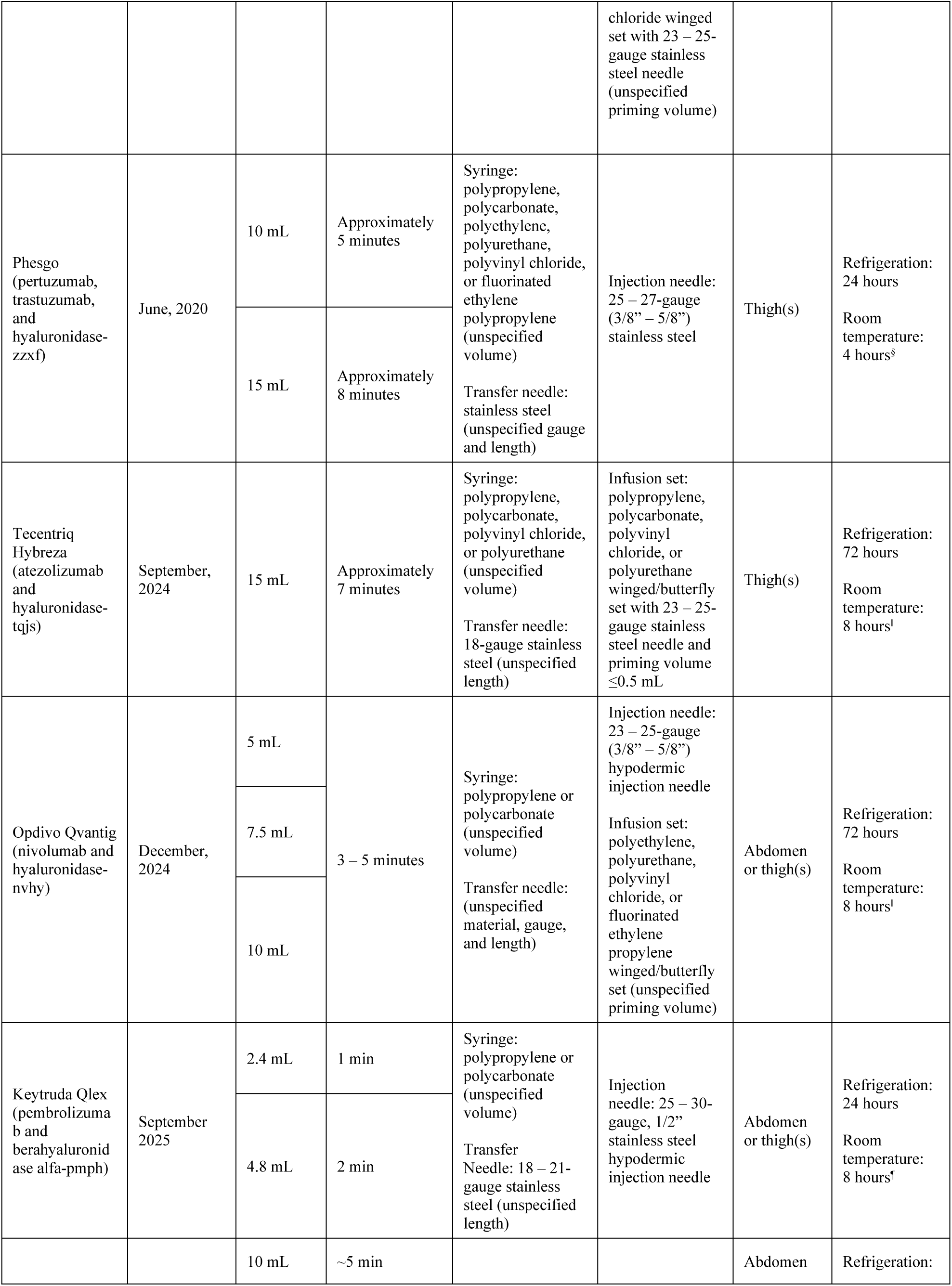
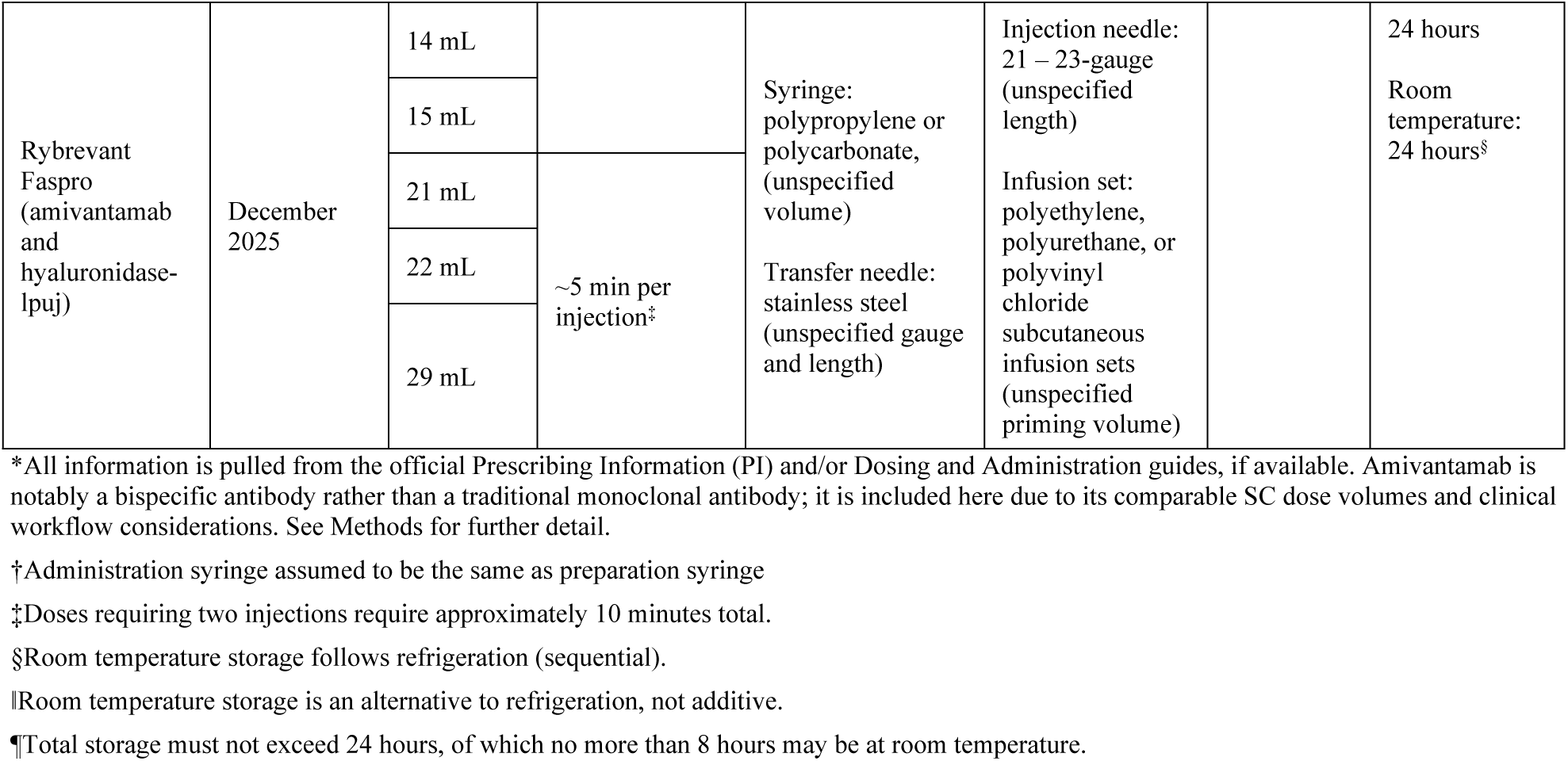
– FDA-Approved SC Oncology Monoclonal Antibodies*.

Despite recommendations in manufacturer prescribing information, there remains some uncertainty in the clinical community about best practices for SC administration of large volumes and little has been published detailing how these products are prepared and administered in real-world use.[37] One recent publication from the Oncology Nursing Society emphasized the lack of evidence-based recommendations for specific techniques or supplies to guide administration of SCOmAbs, urging the nursing community to fill data gaps.[38] The absence of clear guidance in this area and incongruency between manufacturer recommendations and practical HCP workflows have the potential to lead to “workarounds” that could have adverse patient impacts.[39–42] Nurse-driven studies have already suggested deviation from manufacturer administration recommendations for SCOmAbs, exploring the use of other delivery methods to improve nurse and/or patient comfort and minimize risk of nurse musculoskeletal disorders (MSDs) caused by repeated manual syringe injections. Briefly, one study presented alternatives for needle or administration set attachments that could be paired with a syringe pump for administration,[43] two others explored the use of flexible catheters for SC delivery,[44,45] and a fourth demonstrated increased nursing satisfaction and improved comfort with the use of winged infusion sets compared to standard hypodermic needles.[46] Such research appears to be warranted, as ergonomic issues, including increased prevalence of MSDs, have been observed among HCPs performing similar motions or assuming similar postures to those required for manual syringe injections,[47–50] and have the potential to result in mental health issues, diminished productivity, and lower quality of life in those affected.[51–53] Moreover, with the recent approval of SC formulations of widely-used PD-1/L1 inhibitors,[7] advancing SC formulations of other immune checkpoint inhibitors,[54,55] and substantial development of large-volume (>2 mL) SC (LVSC) anti-cancer biopharmaceuticals overall,[56] risk of these complications may increase as SCOmAbs expand to more tumor types and represent greater proportions of administrations.

Augmenting or even replacing manual administration with more automatic delivery devices represents one potential way to offset the practical challenges associated with more frequent SC administrations. One such device, the MyDose single-use injection device, has already been extensively studied for in-clinic delivery of SC trastuzumab,[57–61] although was not commercialized due to perceived lack of benefit without approval for patient self-administration at home.[62] Similarly, other administration devices, including alternative on-body delivery systems (OBDSs) and syringe pumps are also undergoing investigation for administration of SCOmAbs.[63–68] Moreover, Phesgo has already received FDA approval for home administration[69], several other SCOmAbs have already been evaluated or are being actively evaluated for at-home use,[70–74] and the availability of user-friendly delivery devices may represent a critical enabler for this transition.[75,76] However, to date, it is unclear how any such device would fit into current oncology clinical practice, whether as an in-clinic solution or a gateway to home administration, nor what would be required to facilitate safe transition to the home at scale.

To better understand real-world use of SCOmAbs and potential unmet needs, we conducted an exploratory study of current HCP and patient practices with these products. The overarching objectives of this research were to (1) characterize current practice and experience with SCOmAbs delivered in clinic; (2) identify existing pain points with the current SC oncology paradigm from both patient and HCP perspectives; (3) examine how a delivery device could potentially fit current in-clinic workflows; (4) project the impact of increased frequency of SCOmAb injections on workload; and (5) explore patient and HCP perspectives on home administration of SCOmAbs.

## Materials and methods

### Study design

This was a US-based, semi-structured, non-interventional, qualitative study of HCP and patient experiences with SCOmAbs delivered in clinic. Included participants were enrolled into one of three arms: patients, nurses, or pharmacists/pharmacy technicians according to the eligibility criteria described in the subsequent section. Study sessions consisted of 30–60-minute, one-on-one, in-depth interviews (IDIs) conducted between June 21 and July 8, 2024, either in person or by video, depending on participant geography. IDIs were focused on HCP and patient practices, pain points, and behavior related to the preparation and administration of SCOmAbs in outpatient infusion centers and the potential future expansion of SCOmAb delivery, both in terms of frequency and the prospect of home administration. For the purposes of interview efficiency, participants were asked to focus their responses on their current SCOmAb treatment (in the patient arm) or the SCOmAb they were most familiar with of three provided options (in the nurse and pharmacist/pharmacy technician arms). Similarly, topics that have already been thoroughly characterized elsewhere, such as current practice and experience with IV products, precise comparisons of time requirements for SCOmAbs versus their IV alternatives, and overall preference for SC versus IV administration, were not included prospectively in IDIs but rather captured opportunistically only if volunteered by participants.

The breadth of included discussion topics for each study arm linked to the overarching study objectives is provided in Table 2. Discussion topics were deliberately overlapped between study arms to utilize the full sample and gain diverse perspectives on the current workflow and future considerations. Moreover, participants were enrolled from a variety of geographies to capture any regional difference in practices, and no participants came from the same facility. As a result, responses were considered independent rather than dyadic, with no assumed relationship between the practices reported among study arms. Instead, independent responses from each arm were aggregated together to create a comprehensive view of SCOmAb workflow, with portions of the sample providing responses to the discussion topics that were within their purview. For example, all participants were asked about where SCOmAbs are prepared, with the nurse and pharmacy arms responding with their respective facility’s practices and the patient arm responding with their observations during in-clinic visits (e.g., medications prepared at bedside or elsewhere). In contrast, only the nurse and patient arms were asked about administration techniques and locations, as these topics are outside the scope of pharmacy practice.

**Table 2.** – Overview of Study Objectives and Associated Discussion Topics.

| Study Objective | Discussion Topics for Each Study Arm |  |  |
| --- | --- | --- | --- |
|  | Arm 1: Patients<br>(n=10) | Arm 2: Nurses<br>(n=10) | Arm 3: Pharmacy Staff<br>(n=10) |
| 1. Characterize current practice and experience with SComAbs delivered in clinic | Scheduling and logistics of in-clinic appointments<br>Supplies and processes to receive a dose<br>Preparation before and recovery after an injection | Scheduling and ordering of medication<br>Supplies and processes to administer a dose<br>Preparation and transport of the medication between areas of the infusion clinic | Scheduling and ordering of medication<br>Supplies and processes to prepare a dose in the hood<br>Preparation and transport of the medication between areas of the infusion clinic |
| 2. Identify existing pain points with the current SC oncology paradigm from both patient and HCP perspectives | Pain points faced when receiving the injection<br>Actions taken to improve their experience | Pain points faced while administering the injection<br>Actions taken to improve their experience | Pain points faced while preparing the injection<br>Actions taken to improve their experience |
| 3. Examine how a delivery device would potentially fit current in-clinic workflows | Not assessed | Impact of PFS on the administration workflow<br>Impact of delivery device in administration workflow | Impact of PFS on the preparation workflow<br>Impact of delivery device in preparation workflow |
| 4. Project the impact of increased frequency of SComAb injections on workload | Not assessed | Impact on workflows and ergonomics for nurses<br>Impact on staffing and scheduling in the facility | Impact on workflows and ergonomics for pharmacy<br>Impact on staffing and scheduling in the facility |
| 5. Explore patient and HCP perspectives on home administration of SComAbs | Current experience of patient self-administering<br>Benefits and concerns of self-administering at home | Support for and any concerns about patients self-administering at home | Not assessed |

Prior to enrollment, all participants completed a written informed consent form outlining the observational nature of the study, the requirements for participation, the intent to publish de-identified study data in a scientific journal, and the steps taken to protect their privacy throughout. Participants were informed that they could withdraw at any time without consequence, including if they expressed discomfort with any study requirement or having their de-identified data shared for scientific purposes. All participants were compensated financially for their time. Participants were blinded to the identity of the pharmaceutical sponsor, and the sponsor was blinded to participant identities. During the IDIs, participants were first asked to describe their current workflow with SCOmAbs before any further discussion or projective exercises took place in order to reduce the risk of biasing their description of current process and experience. All study procedures were conducted in accordance with the 1964 Declaration of Helsinki and its subsequent revisions. This was a non-interventional, observational study in which all data were collected through interview procedures, recorded using non-identifiable participant codes, and reported in aggregate or with de-identified quotations. Both Matchstick LLC and AstraZeneca independently determined that this research met the criteria for exempt research under United States 45 CFR 46.104(d)(2) and did not require Institutional Review Board approval.

### Participant eligibility

Participant recruiting was performed throughout the United States using commercially available, nationwide panels of HCPs and patients with chronic disease. Enrolled participants were ≥18 years of age and had experience with SCOmAbs as a patient, nurse, or pharmacist/pharmacy technician at the time of study enrollment, as outlined above. Pharmacists and pharmacy technicians were recruited separately but later combined into a single group to account for variability in medication preparation practices across different facilities.

Eligible patient participants had a formal diagnosis of multiple myeloma, diffuse large B-cell lymphoma, chronic lymphocytic leukemia, early-stage breast cancer, or metastatic breast cancer, and were currently receiving SC injections with at least one of Darzalex Faspro (daratumumab and hyaluronidase-fihj), Rituxan Hycela (rituximab and hyaluronidase human), Herceptin Hylecta (trastuzumab and hyaluronidase-oysk), or Phesgo (pertuzumab, trastuzumab, and hyaluronidase-zzxf). Tecentriq Hybreza (atezolizumab and hyaluronidase-tqjs), Opdivo Qvantig (nivolumab and hyaluronidase-nvhy), Keytruda Qlex (pembrolizumab and berahyaluronidase alfa-pmph), and Rybrevant Faspro (amivantamab and hyaluronidase-lpuj) were not included, as none of these products were approved at the time of study initiation. Notably, however, these more recently approved products have dose volumes similar to or exceeding those of the SCOmAbs included in this study: Tecentriq Hybreza, Opdivo Qvantig, and Rybrevant Faspro are administered in 15 mL, up to 10 mL, and up to 29 mL dose volumes, respectively, while Keytruda Qlex is administered in up to a 4.8 mL dose volume. Although the term “SCOmAb” is used throughout this manuscript, it should be noted that amivantamab is a bispecific antibody rather than a traditional monoclonal antibody, and is included in the broader discussion because its large SC dose volumes are likely to present similar clinical workflow considerations as the SC monoclonal antibodies studied here. Other SC oncology antibodies administered in smaller volumes, such as recently approved bispecific T-cell engagers, are not addressed here, as they are not expected to present comparable LVSC administration challenges.

Eligible nurse participants had recent experience (within the last month) administering at least one of the four SCOmAbs listed above in an outpatient infusion center. Eligible pharmacists and pharmacy technician participants also had recent experience (within the last month) preparing at least one of the four SCOmAbs in an outpatient infusion center pharmacy. Participants in any group were excluded if they did not have a reliable internet connection and access to a computer, as these were required for study recruitment and/or study sessions for remote participants. Participants were also excluded if they or an immediate family member worked for a pharmaceutical company or a market research firm to reduce the potential for bias.

### Study stimuli

Discussion topics related to potential delivery device fit with current in-clinic workflows and patient perspectives on self-administration of SCOmAbs at home were supported with simple study stimuli to guide participants with appropriate context. Exploration of potential delivery device fit included the prospect of using a prefilled syringe (PFS) or portable infusion pump to deliver SCOmAbs. To facilitate this discussion, pharmacist/pharmacy technician and nurse participants were shown visual stimuli to illustrate representative PFS and portable infusion pump configurations. For the PFS stimulus, HCPs were shown an investigator-taken photo depicting a packaged Hizentra^®^ (immune globulin subcutaneous [human], 20% liquid) 20 mL PFS aligned alongside a packaged 20 mL polypropylene syringe (Becton Dickinson, Franklin Lakes, NJ, USA) to represent the approximate size and appearance of a volume-matched PFS presentation compared to existing syringes. For the portable infusion pump stimulus, HCPs were shown side-by-side, publicly available images of three on-market portable infusion pump alternatives to illustrate a range of potential pump appearances: Freedom60^®^ (KORU Medical Systems, Chester, NY, USA), Crono S-PID 50 (Canè Medical Technology, Turin, Italy), and Curlin^®^ 4000 CMS^™^ (Moog Medical, Salt Lake City, UT, USA).

Notably, all visual stimuli were introduced to participants purely as examples to clarify the terms “prefilled syringe” and “portable infusion pump” and avoid confusion with conventional syringes and hospital-grade volumetric infusion pumps, respectively. None of the examples were intimated as specific solutions. Similarly, to facilitate exploration of patient perspectives on SCOmAb self-administration at home, patient participants were shown a generic schematic of a portable infusion pump and the components it could potentially encompass (i.e., pump, medication container, tubing set, needle) in order to minimize the risk of confusion associated with particular pump designs. Again, this was intended to help patients conceptualize what self-administration could entail and distinguish administration with a portable delivery device from the methods they were likely already familiar with, namely manual syringe delivery and stationary infusion pump delivery (i.e., a hospital-grade volumetric pump placed on an IV pole).

### Data collection and analysis

Interview sessions were recorded for video and audio, transcribed, and coded by the study moderator (MA). Data was collected through moderator questioning and passive observations. Initial coding was executed as participants completed their interview session to develop an opening codebook, followed by an iterative process of reviewing and refining coding sequences to develop mutually exclusive themes. Thematic saturation was assessed iteratively during data collection and was considered achieved when subsequent interviews within each study arm yielded no substantively new themes. Common workflow elements and facility practices were recorded and coded based on each participant’s reported current practices and pain points to encompass those related to preparation, administration, and receipt of the injection. Quantitative analysis was used to communicate the range of responses to numeric-based questions, such as injection time and total time in clinic. However, due to the relatively small sample sizes and lack of uniform workflow steps for comparison between study arms, no statistical analyses were performed. The study moderator (MA) had no prior relationship with any participant and no clinical background, which helped reduce the potential for leading questions or assumptions about clinical workflows during interviews. Participants were informed that the research was being conducted by Matchstick, an independent research group, but were not told about the study’s funding relationship with AstraZeneca or any specific product development objectives. The broader research team included members with consulting relationships in the pharmaceutical and device industries (CF, MC) and employees of AstraZeneca (JW, AR, SB). To mitigate potential bias from these affiliations, analysis and interpretation were led by the researchers who conducted the interviews and coding (MA, CF), with all authors reviewing findings during manuscript preparation.

## Results

A total of 30 participants were successfully recruited into this study, with 10 enrolled into each study arm. All participants completed every discussion topic outlined in Table 2 with two exceptions based on unique practices. One patient participant was discovered to be currently performing her own injections at-home rather than receiving them in the clinic and was therefore excluded from some topics related to in-clinic administration. Similarly, one nurse participant exclusively administered SCOmAb injections with a syringe pump attachment and was therefore excluded from some topics related to manual push administration. Instead, residual interview time with these two participants was spent opportunistically exploring their unique practices. Participant demographics for all study arms, disease- and treatment-specific characteristics for the patient arm, and chosen SCOmAb example for the nurse and pharmacy staff arms are presented in Table 3, Table 4, and Table 5, respectively. Additional participant-level detail, including diagnosis, current SCOmAb, and prior IV formulation experience for each patient, is provided in S2 Table; corresponding detail for nurse and pharmacy staff participants is provided in S3 Table. Participant responses to the discussion topics are provided in the subsequent sections, organized by the study objectives and their associated subtopics.

**Table 3.**
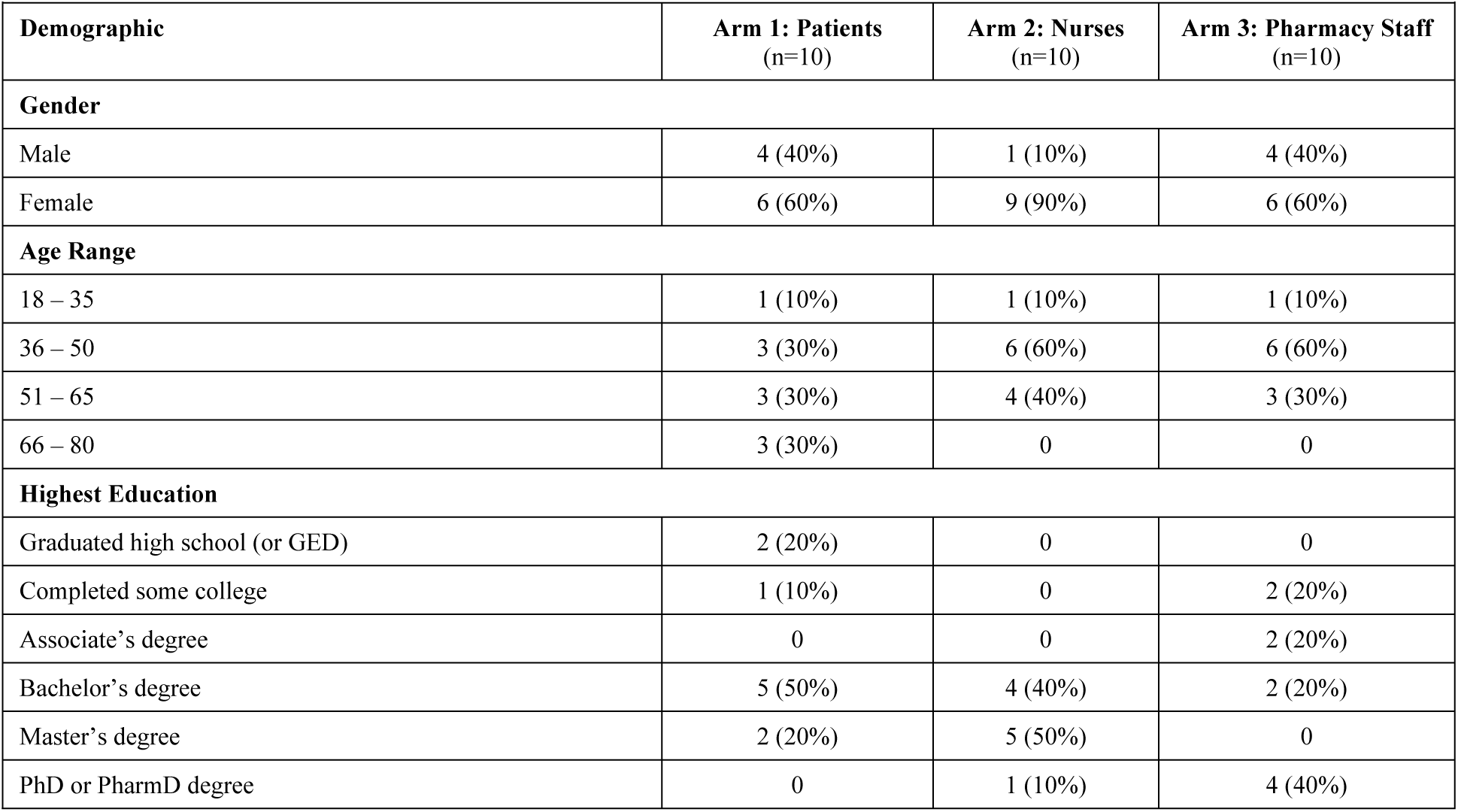
– Participant Demographics.

| Demographic | Arm 1: Patients<br>(n=10) | Arm 2: Nurses<br>(n=10) | Arm 3: Pharmacy Staff<br>(n=10) |
| --- | --- | --- | --- |
| **Gender** |  |  |  |
| Male | 4 (40%) | 1 (10%) | 4 (40%) |
| Female | 6 (60%) | 9 (90%) | 6 (60%) |
| **Age Range** |  |  |  |
| 18 – 35 | 1 (10%) | 1 (10%) | 1 (10%) |
| 36 – 50 | 3 (30%) | 6 (60%) | 6 (60%) |
| 51 – 65 | 3 (30%) | 4 (40%) | 3 (30%) |
| 66 – 80 | 3 (30%) | 0 | 0 |
| **Highest Education** |  |  |  |
| Graduated high school (or GED) | 2 (20%) | 0 | 0 |
| Completed some college | 1 (10%) | 0 | 2 (20%) |
| Associate's degree | 0 | 0 | 2 (20%) |
| Bachelor's degree | 5 (50%) | 4 (40%) | 2 (20%) |
| Master's degree | 2 (20%) | 5 (50%) | 0 |
| PhD or PharmD degree | 0 | 1 (10%) | 4 (40%) |

**Table 4.** – Patient Disease- and Treatment-Specific Characteristics.

|  |  |
| --- | --- |
| **Primary Disease State** |  |
| Multiple myeloma | 4 (40%) |
| Metastatic breast cancer | 3 (30%) |
| Early breast cancer | 1 (10%) |
| Diffuse large B-cell lymphoma | 1 (10%) |
| Chronic lymphocytic leukemia | 1 (10%) |
| **Current SComAb** |  |
| Darzalex Faspro | 4 (40%) |
| Herceptin Hylecta | 3 (30%) |
| Rituxan Hycela | 2 (20%) |
| Phesgo | 1 (10%) |
| **Prior Experience with IV Formulation of Current SComAb** |  |
| Yes | 6 (60%) |
| No | 4 (40%) |

**Table 5.** – Nurse and Pharmacy Staff Facility Characteristics.

| Table 5 – Nurse and Pharmacy Staff Facility Characteristics |  |  |  |
| --- | --- | --- | --- |
|  | Arm 2: Nurses<br>(n=10) | Arm 3: Pharmacy Staff |  |
|  |  | Pharmacists (n=5) | Pharmacy Techs (n=5) |
| SCOmAb Chosen for Discussion |  |  |  |
| Darzalex Faspro | 4 (40%) | 4 (80%) | 4 (80%) |
| Phesgo | 4 (40%) | 1 (20%) | 0 |
| Rituxan Hycela | 2 (20%) | 0 | 1 (20%) |
| Approximate Facility Size (Number of Infusion Chairs) |  |  |  |
| Small (< 20) | 3 (30%) | N/A * |  |
| Medium (20 – 50) | 3 (30%) |  |  |
| Large (> 50) | 4 (40%) |  |  |
| Frequency of SCOMAb Dose Preparation/Administration |  |  |  |
| < 1 dose daily | 0 | 0 | 1 (20%) |
| 1 – 10 doses daily | 3 (30%) | 2 (40%) | 2 (40%) |
| 11 – 20 doses daily | 2 (20%) | 0 | 1 (20%) |
| 21 – 50 doses daily | 1 (10%) | 2 (40%) | 0 |
| Unknown (could not approximate) | 4 (40%) | 1 (20%) | 1 (20%) |

\*Not collected for pharmacy sample, as considered out of pharmacy purview.

### Characterize current practice and experience with SCOmAbs delivered in clinic

#### Aggregate workflow and facility variability

An aggregate workflow summary comprising participant practices with associated counts is provided in Fig 1. Based on participant responses, the workflow was subdivided into sections according to which study arm is involved, when practices occur relative to others, and how practices vary between facilities in our sample. Overall, practices generally mirrored those of typical clinical workflows for parenteral medications, such as IV infusions (Fig 1, “Expected Behavior”). However, there was also considerable variability in SCOmAb preparation and administration processes between facilities. Most notably, three main deviations from expected IV infusion workflows and/or specifications provided in SCOmAb product labeling were observed (Fig 1, “Deviation”): (1) 7/30 participants (5 patients and 2 nurses) reported that nurses prepare the medication at the patient’s bedside (i.e., attach a needle to a conventional syringe, insert the needle into the medication vial, draw the medication into the syringe, and adjust to the required dose); (2) 1/30 participant (1 patient) reported that she self-administers the medication at home; and (3) 3/19 participants (3 nurses; sample excludes the 1 patient who self-administers and all 10 pharmacists/pharmacy technicians, as they did not have visibility to administration practices) reported that a syringe pump module attached to an infusion pump is used to automate medication administration. These deviations and overall participant-reported practices with SCOmAbs are detailed in the following sections, organized according to sections A-E in Fig 1.

**Fig 1.**
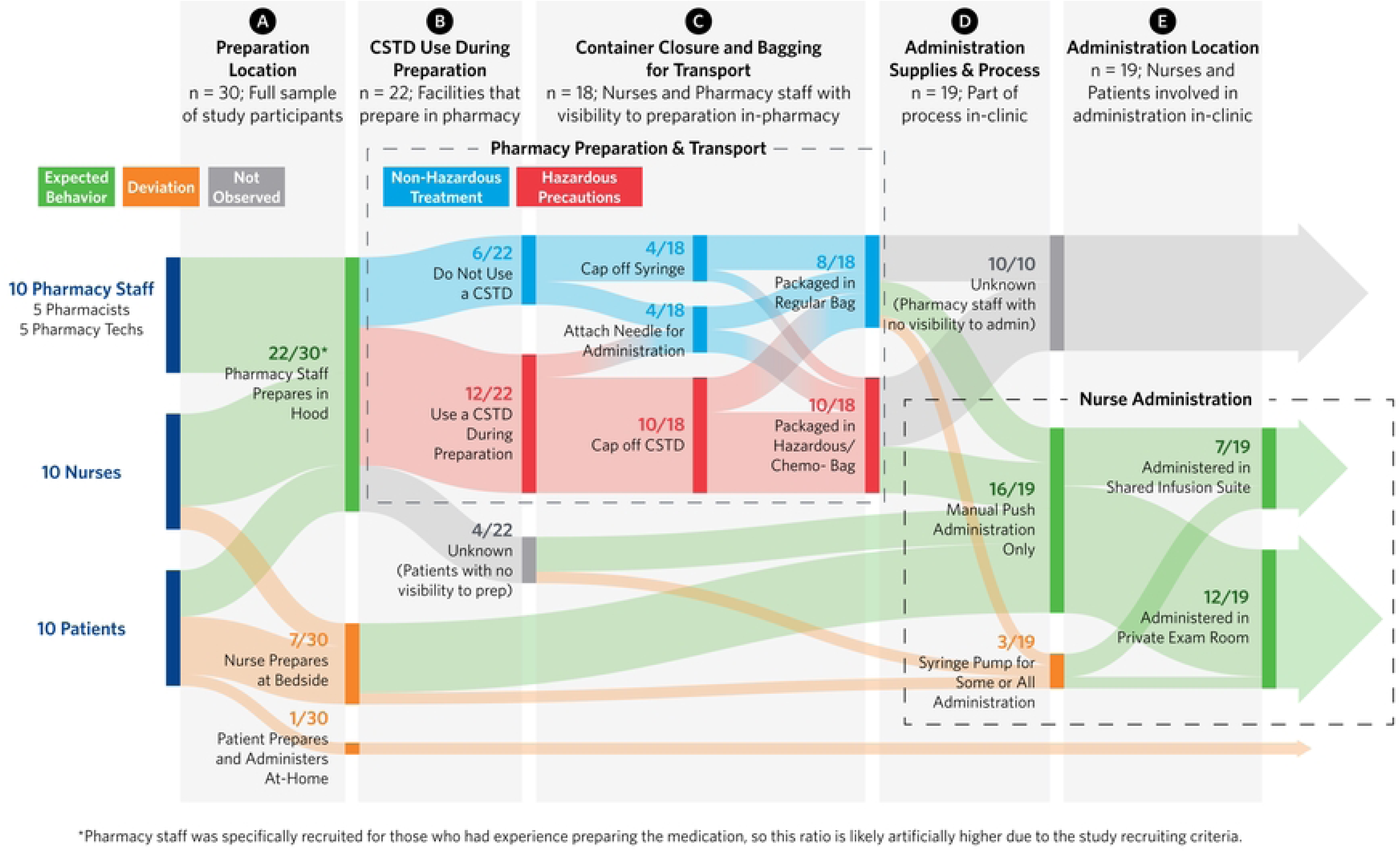
Facility practices for preparation and administration of SC oncology injections. (n = 30 participants including the 10 patients, 10 nurses, and 10 pharmacy staff)

#### Preparation location within the facility

Participant reports of where (i.e., the location within their facility) SCOmAbs are prepared are summarized in Fig 1, section A. Across all three study arms, most participants in our sample (22/30) reported that SCOmAb preparation occurs in their facility’s pharmacy compounding areas, similar to where most IV medications are prepared. As experience with SCOmAb preparation was a recruiting criterion for the pharmacy staff arm, all participants in this group expectedly reported that these agents are prepared in pharmacy, specifically in compounding hoods (e.g., laminar airflow workbenches, isolators, or biological safety cabinets depending on individual facility infrastructure), with minimal variation in their preparation process compared to typical IV workflows. Pharmacists and technicians emphasized that these preparations are generally simpler compared to many IV medications, as they require a single and straightforward draw of a fixed dose. On this topic, one pharmacist commented:

> *“Darzalex Faspro is a straight draw. There’s no dilution, there’s really nothing. It’s pretty consistent.” **–*** **P5PR**

In contrast, preparation location was more variable in the nurse and patient arms. While most nurses (8/10) still reported that preparation is performed in pharmacy, the remaining small portion (2/10) described that they perform SCOmAb preparation themselves at the bedside. In the patient group, half of participants (5/10) reported observing their nurses performing the preparation, with the remainder either reporting that their nurse receives an already-filled syringe from the pharmacy (4/10) or in one unique case (mentioned above and discussed separately below), they prepare the medication themselves at home (1/10). One nurse in our sample who performs beside preparation emphasized that this process has the potential for both operational and patient care benefits:

> *“They [Pharmacy] love it when they can give it to the nurses, and I don’t mind drawing it up. I feel like it’s safe and it’s not a problem. You can still talk to the patient while you’re getting things ready, and they can watch […] It’s almost like when you go to a restaurant and they’re cooking up front instead of in the back, and you really watch the process. It can be comforting [for the patient].” **–*** **P14NR**

#### CSTD use during preparation

The packaging and handling of completed SCOmAb preparations (i.e., filled syringes) for transport from the pharmacy to the administering nurse is captured in Fig 1, section B, with detail on CSTD use and facility treatment of these medications as hazardous versus non-hazardous. Among participants who reported that SCOmAb preparation occurs in pharmacy, most (12/22) reported that CSTDs are used and only a small portion (6/22) reported they are not (the remaining 4/22 comprised patients with no direct visibility to preparation, and use of CSTDs was therefore unknown). Those participants who do use CSTDs during preparation in the pharmacy (12/22) reported experiencing issues with broken components (3/12), more cumbersome systems that require specific adaptors (3/12), and greater potential to introduce air bubbles into the medication during preparation (1/12). The remaining HCPs that use CSTDs for SCOmAbs (5/12) reported that they have not experienced issues with them.

CSTD use and overall treatment of SCOmAbs as hazardous medications was not always consistent throughout the preparation process among those who reported using them. Some (2/12) pharmacists/technicians and nurses reported that the administration needle is attached onto the CSTD in the hood after preparation, thereby opening the fluid path and compromising the closed system. Others (5/12) reported that filled SCOmAb syringes are not packaged in chemotherapy/hazardous medication bags for transport (implying that the medications inside *are not* hazardous) despite using CSTDs during preparation (implying that the medications being prepared *are* hazardous). Similarly, some (2/6) who reported no CSTD use during preparation still transport or receive SCOmAbs in chemotherapy/hazardous medication bags.

#### Container closure and transport

Additional detail on closing and transporting filled syringes after SCOmAb preparation is presented in Fig 1, section C. Among participants in our sample who reported that SCOmAbs are prepared in pharmacy, most (14/18) stated that filled syringes are capped or otherwise sealed, and it is the administering nurse who is later responsible for selecting and attaching injection needles or tubing sets. However, the remaining (4/18) participants reported that pharmacy staff will attach administration needles or tubing sets to reduce steps and complexity for the administering nurse. One nurse participant specifically mentioned that high staff turnover has impacted her facility’s training burden, and that pharmacy providing ready-to-use syringes simplifies administration steps and reduces error potential for new nurses:

> *“The fact that [the pharmacy] gives us everything prepared works really well, and it saves time for us because we’re so crunched with time. We do have a lot of new staff that didn’t come from oncology backgrounds, so we’re always training, and even when things are fully prepared, they still sometimes don’t know how to handle a lot of things. I do think that it’s very helpful for us with time management, that pharmacy just gives us everything ready to go, and it’s like almost impossible, not impossible, but almost impossible to make a mistake.” **–*** **P19NR**

When pharmacy preparation is complete, participants reported that a variety of methods are used to transport SCOmAbs to administration areas according to facility layout and available infrastructure. Transport practices in our sample included nurse retrieval from pharmacy (9/20), pharmacy delivery to administration areas (6/20), and use of more facilitated methods like dumbwaiters (3/20), automated dispensing cabinets (1/20), or pneumatic tube systems (1/20). All of these methods include some degree of waiting period between when medications are ordered and when they are received for administration. One nurse in our sample reinforced this, stating:

> *“Most of the time we’re just kind of running around to catch up, so we’re always waiting for meds.” **–*** **P16NR**.

#### Administration supplies and process

Practices specific to the SCOmAb administration process (i.e., needle attachment and injection) are shown in Fig 1, section D. While preference for SC vs. IV administration was not a primary interview focus as previously noted, nurses volunteered that SCOmAbs offer unique benefits compared to IV-administered oncology agents, including reduced overall time and greater convenience as a result. One nurse specifically noted that although the SC route requires her to sit with patients longer for the administration itself (i.e., compared to starting an IV pump), this time is more “casual” and offers the opportunity to facilitate more intimate conversations:

> *“It’s definitely a more intimate time with the patient because you are sitting there with them. And I’ve had like amazing conversations, like goals of care conversations with people literally while we’re doing it. And so for me to have like 5 minutes just with them, and really find out how they’re doing and what’s going on and what’s important to them, and leading them into conversations like that, if they’re open. I like doing that. I mean, because when you’re putting in an IV, it’s a lot more technical, really, you know, than when you’re putting a needle into the fat of somebody’s belly. It’s not like as precise. And so there’s a lot, you know, there’s a lot of focus when you’re putting in an IV, and this [SC administration] seems a little bit more casual.” **–*** **P14NR**

In terms of administration supplies, nurses in our sample described that they are limited to the supplies their facilities stock to perform SCOmAb injections, with variation in supply use driven by whether or not CSTDs are used, which needle types are stocked, and whether or not nurses prepare the medications themselves rather than receive filled syringes from pharmacy.

Syringe and needle selection were particular areas of interest for administration supplies, as pharmaceutical manufacturer labeling provides more specific guidance on these compared to other supplies. In our sample, selection of syringe size was typically informed by SCOmAb dose volume and any facility restrictions on maximum syringe fill volumes, with no mention of product labeling. One pharmacist specifically commented:

> *“For subcutaneous, we usually use the syringe closest to whatever the dose is, so 15 [mL dose], we’re using a 20 [mL syringe].” **–*** **P22PR**

Moreover, participants reported that syringe selection is uniformly determined by whoever prepares the medication, and if a pharmacist or technician does so, the administering nurse has no influence over which syringe is used for administration. However, some nurses did report that their pharmacy may select a larger syringe than required to house a dose, with the intent to “leave enough space” for them to manipulate the syringe without risking leakage or plunger dislodgement out the back of the syringe:

> *“So, we have a policy where the [dose] volume needs to be less than 70% of the syringe [volume]…the reason is because we don’t want the volume to be so big in the syringe, where, you know, there might be an accidental like [plunger] coming out from [the back of] the syringe. So as long as it’s less than 70%, then we kind of go to the nearest syringe size based on that.” **–*** **P5PR**

Restrictions on maximum SC injection volumes per injection site varied between facilities in our sample, although only one facility imposed a limit below 15 mL (the maximum dose volume of studied SCOmAbs). This facility instead opted for maximum of 7-8 mL per site, therefore requiring two injections for any 15 mL dose. Patient and nurse experiences related to dose splitting to multiple injection sites are summarized in Table 6. Overall, most nurses (6/10) and patients (7/10) reported that only one injection site is used, unless the facility protocols prevent it (1/10 nurse report), there is an issue with the first injection site that forces use of an additional injection site (1/10 nurse report and 2/10 patient reports), or a patient makes a specific request to split the dose (1/10 nurse report). With one exception, all patients and nurses reported that the full dose is still prepared in one syringe regardless of these circumstances, and is only proactively split between two syringes at the one facility in our sample that imposes SC volume restrictions.

**Table 6.** – Nurse and Patient Experience with Dose Splitting Between Multiple Sites.

| Dose Splitting Behavior | Count for Nurses (n=10) | Count for Patients (n=10) | Quote Identifier | Representative Quote |
| --- | --- | --- | --- | --- |
| Never splits the dose | 6 (60%) | 7 (70%) |  |  |
| Always with anything above 7 – 8 mL as a facility-wide practice | 1 (10%) |  | Q1 | “You don’t want to create like any swelling. And edema injecting, injecting a large volume over a short period of time. Then you start thinking about the absorption and the discomfort, the pain.” – **P7NR** |
| Sometimes to reduce discomfort for select patients | 1 (10%) | 2 (20%) | Q2 | “What drives that is, having a patient really complain about the discomfort from the single injection, and then going and speaking to the pharmacist about splitting the dose in into 2 separate syringes.” – **P28NR** |
| Only due to an issue in the 1<sup>st</sup> site that forces a 2<sup>nd</sup> site | 1 (10%) | 1 (10%) | Q3 | “It was the patient. He couldn’t tolerate it. It was very painful. But he did have an injection in that site before, so it was a little hard, and it was bruised. But, you know, I was running out of sites to rotate and based on, you know, the documentation, that was the next site where we had to go. [...] But, you know, you have to go by what the patient says, and he’s in pain, he’s uncomfortable. I wasn’t going to continue, so I rotated the site. That only happened once.” – **P19NR** |
| Only for one specific patient who requests it | 1 (10%) |  | Q4 | “She’s [the patient] a physician herself. She’s super fit and thin and very explicit about what she wants, and so it’s fine with me. She wants it split into two, so I use two [10 mL syringes] and did a half and half [...] she would have it bilateral in her lower abdomen. That’s what she prefers.” – **P14NR** |

Those injecting the dose from one syringe into two sites reported that they do so using separate administration needles, and no instances of needle reuse were reported.

Needle choice was also influenced by facility practice in our sample, although compared to syringe choice, individual nurses expressed that they had more autonomy to choose between available needles. Many (6/10) nurses in our sample reported that they follow consistent, facility-wide practices for needle selection, but the remainder reported nurse-specific variation according to their own preferences, with more preferring butterfly needles to straight needles (5/10 versus 4/10 nurse reports) if available:

> *“It [the butterfly needle] stays beautifully. I put it [the butterfly needle] all the way in and I have like a free hand now to basically time myself as I’m pushing the syringe. I want to say, it’s like less resistance too and I feel like the patient also feels a little bit less nervous because you’re like there, but you’re not like touching them directly.” **–*** **P16NR**

A summary of needle type and gauge selection according to both nurse and patient participants is captured in Table 7. Most nurse participants reported that they primarily use either a 23- or 25-gauge straight SC needle (2/10 nurse reports and 2/10 nurse reports, respectively) or a 23-gauge butterfly needle set (5/10 nurse reports). The remaining nurse (1/10) described a unique practice that involves repurposing a 24-gauge IV catheter to facilitate SCOmAb administration with a syringe pump. Notably, these selected needle types and/or gauges were not always consistent with those recommended in the corresponding products’ prescribing information (Table 7). Regardless, nurses expressed that they prefer to use the largest needle available to reduce resistance in the system and speed up the injection, while still maintaining patient comfort as much as possible. One nurse shared her facility’s reasoning for switching to larger needles even if it meant stocking an atypical product:

> *“We made that change to a little bit of a larger needle [25-gauge] based on the feedback that pushing Phesgo through a 27 [gauge] felt really, really difficult and much slower. People felt it just took longer and wasn’t moving as quickly as they wanted it to. […] I actually had to bring new stock to the units, because we don’t normally carry a 25-gauge needle for subcutaneous.” **–*** **P4NR**

**Table 7.** – Needle Types Used by Participants.

|  | Count for Patients<br>(n=10)* | Count for Nurses<br>(n=10) |  |  | Count for Nurses<br>(subgroups stratified by presence/absence<br>of selected needle type/gauge in<br>corresponding product PI) |  |
| --- | --- | --- | --- | --- | --- | --- |
|  | Needle Type | Needle Type | Needle Gauge |  | On-Label | Off-Label |
| Straight Needle | 7 (70%) | 4 (40%) | 25-gauge | 2 (20%) | 2 (100%) | - |
|  |  |  | 23-gauge | 2 (20%) | 1 (50%) | 1 (50%) |
| Butterfly Needle | 3 (30%) | 5 (50%) | 23-gauge | 5 (50%) | 2 (40%) | 3 (60%) |
| IV Catheter |  | 1 (10%) | 24-gauge | 1 (10%) | - | 1 (100%) |

\*The patient who self-administered at home did so using a straight needle

#### Administration location within the facility

Practices specific to the SCOmAb administration location (i.e., where injections are performed within a facility) are shown in Fig 1, section E. When patients and nurses were asked where in their facility SCOmAbs are administered (out of scope for pharmacists/pharmacy technicians), answers varied between one of two locations. Most (7/10) nurses reported using shared areas within their infusion suite and pulling a curtain around the patient for privacy during the injection, while others (3/10) use private rooms, noting that this is particularly necessary for patients who must remove clothing for thigh injections. Conversely, most (5/9) patients in our sample reported always receiving their injection in a private room, with others either always using a shared area with curtains drawn (2/9) or variably using either one of these two locations depending on the visit (2/9).

### Identify existing pain points with the current SC oncology paradigm from both patient and HCP perspectives

#### Injection timing and ergonomics

Nurse heuristics for SCOmAb administration, including injection speed relative to prescribing information recommendations and timekeeping methods, are captured in Table 8. Many (6/9) nurses in our sample reported that they rely on a wristwatch (5/9 reports) or wall clock (1/9 reports) to passively keep time while they watch the syringe plunger move relative to the syringe graduations, comparing the volume remaining to the time remaining to gauge their progress. Others (3/9) expressed that they simply use muscle memory to control the plunger speed without any time reference. None of these techniques were described as particularly precise, with one nurse commenting:

> *“I like to just watch that I’m going down in volume and I just assume if I’m doing 2 to 3 mL every couple of minutes, I pretty much hit the mark, but no real science to it.” **–*** **P4NR.**

**Table 8.**
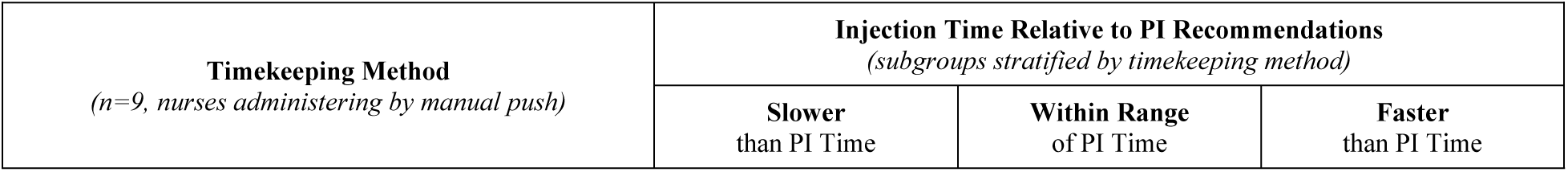
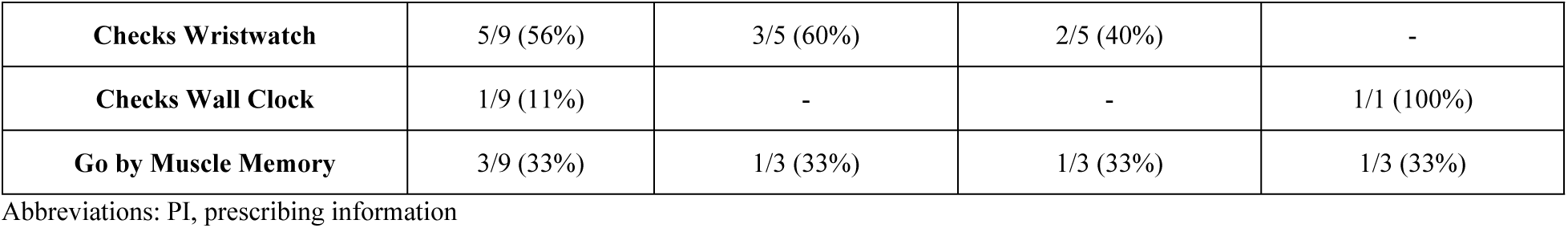
– Timekeeping and Injection Times Compared to PI Recommendations.

|  |  |  |  |
| --- | --- | --- | --- |
| **Timekeeping Method**<br>(n=9, nurses administering by manual push) | **Injection Time Relative to PI Recommendations**<br>(subgroups stratified by timekeeping method) |  |  |
|  | **Slower**<br>than PI Time | **Within Range**<br>of PI Time | **Faster**<br>than PI Time |

|  |  |  |  |  |
| --- | --- | --- | --- | --- |
| **Checks Wristwatch** | 5/9 (56%) | 3/5 (60%) | 2/5 (40%) | - |
| **Checks Wall Clock** | 1/9 (11%) | - | - | 1/1 (100%) |
| **Go by Muscle Memory** | 3/9 (33%) | 1/3 (33%) | 1/3 (33%) | 1/3 (33%) |

Abbreviations: PI, prescribing information

The majority (6/9) of nurses also described that they inject either more slowly (4/9 reports) or more rapidly (2/9 reports) than SCOmAb prescribing information recommendations, citing that they rely on guidance by word of mouth within their facilities rather than consulting the product prescribing information directly. Those who inject more slowly expressed that they do so deliberately to reduce discomfort for the patient, while those who inject more rapidly do so because it is considered acceptable in their facility. Independent of their chosen injection speed, all nurses (10/10) reported that they will address any patient discomfort by pausing the injection and waiting until the patient is ready to continue.

The ergonomics of the injection process, presence/absence of physical discomfort during administration, and specific syringe push techniques used by nurses in our sample are summarized in Table 9. Overall, nurses’ chosen administration positions and motions were highly influenced by their experience with other medications and personal preferences. Most (8/9) nurses in our sample reported that they take the time to set up a comfortable seated position for the duration of the injection, which typically involves setting up a chair next to the patient and adjusting the bed height accordingly. These nurses described that this positioning affords them much more stability compared to standing and leaning over the patient, although some did mention that their colleagues choose to stand instead. One nurse described the impact of uncomfortable positioning on their ability to properly stabilize the injection site:

> *“I used to do it standing up because that’s how everyone else did it. And then I said, this is terrible. It was hurting my back, and I’m kind of leaning over, and I didn’t feel like I had good placement for the patient also. So then I said I need to start making myself comfortable so I could administer this better.” **–*** **P19NR**

**Table 9.** – Ergonomics of Manual Push Technique.

| **Physical Discomfort for the Nurse** | **Physical Position of the Nurse** | **Manual Push Technique** |
| --- | --- | --- |

| (n=9, nurses administering by manual push) |  | (subgroups stratified by presence/absence of physical discomfort) |  | (subgroups stratified by presence/absence of physical discomfort) |  |
| --- | --- | --- | --- | --- | --- |
|  |  | Standing | Sitting | Continuous | Pulsatile |
| Yes | 5/9 (56%) | - | 5/5 (100%) | 4/5 (80%) | 1/5 (20%) |
| No | 4/9 (44%) | 1/4 (25%) | 3/4 (75%) | 3/4 (75%) | 1/4 (25%) |

Nurses reported that they maintain their chosen position throughout the administration, which had some implications for the selected administration needle. For example, nurses who use a butterfly needle reported that they do not use adhesive (e.g., Tegaderm) to secure the site, as they do not change positions during the injection in ways that could lead to needle dislodgment. Moreover, only a few (2/5) of these nurses reported that they stabilize the butterfly needle assembly at the site with their free hand, with the majority (3/5) instead relying on the patient staying still to keep the needle in place during administration.

For the injection itself, most (7/9) nurses reported that they use a continuous push (i.e., constant pressure on the syringe plunger) to administer, while a few (2/9) use a pulsatile method (i.e., sporadic periods of pressure on the syringe plunger). In either case, the chosen push technique was reported to be mostly related to current practice with other medications (e.g., those given by IV push) rather than ergonomics. Still, many (5/9) nurses reported experiencing some degree of physical discomfort and/or pain associated with the injection process even after finding a comfortable and stable position. The types of nurse-reported physical discomfort and their associated descriptions are provided in Table 10. Nurses most frequently reported fatigue in the hand or resting arm (3 reports and 2 reports, respectively) and difficulty maintaining stability at the needle site (2 reports).

**Table 10.** – Nurse Experience with Discomfort During Manual Push Administration *(n=5 nurses who expressed discomfort with manual push administration)*

| Type of Discomfort<br>(select all that apply) | Count for Nurses<br>(n=5) | Quote Identifier | Representative Quote |
| --- | --- | --- | --- |
| Fatigue in the hand or arm regardless of position | 3/5 (60%) | Q5 | “My hand does at times get a little cramped.” – P28NR |
| Difficulty keeping syringe stable without second hand resting on the barrel | 2/5 (40%) | Q6 | “I always feel very far from the injection site which always makes me question like how good my needle control is if that makes sense.” – P4NR |
| Fatigue in the arm if elbows aren't resting on a surface | 2/5 (40%) | Q7 | "If I have my arm set somewhere my arm doesn't get tired." – P4NR |

#### Impact of volume and viscosity

As reflected in Table 5, all HCP participants in this study selected SCOmAbs with comparatively large SC dose volumes (10-15 mL range) for discussion, with the result that the workflow challenges described above pertain primarily to these larger-volume products. However, to explore whether a smaller volume would alter these perspectives, participants were also asked how a 5 mL dose would impact their workflow and pain points as a point of comparison. For participants in the pharmacist/pharmacy technician group, the difference between a 5 mL and 15 mL dose size had no reported impact on their process. In contrast, many (6/10) nurse participants reported several differences between smaller and larger dose volumes, including: smaller welts produced at the injection site and less associated pain for patients (4/6 reports), faster injections that are physically easier to manually administer (3/6 reports), smaller syringes that can be used to deliver the full dose of medication (2/6 reports), and a greater propensity for nurses to prepare the medication themselves instead of having the pharmacy prepare it (1/6 reports).

Compared to medications with water-like viscosities, the SCOmAbs assessed in this study are more viscous (S1 Table). Interestingly, however, pharmacists/pharmacy technicians in our sample did not report that they perceived SCOmAbs as particularly viscous, as they typically employ large gauge needles (e.g., 16-gauge, 18-gauge) to manipulate them during preparation. Nurses, in comparison, reported that the combination of comparatively viscous SCOmAb drug products and the smaller gauge needles used for administration can create challenges, including difficulties pushing the plunger (7/10 reports), clogging in the needle that requires needle replacement (3/10 reports), and added time requirements to bring the medication to room temperature to reduce viscosity (2/10 reports).

#### Patient experience on injection day

Among patients in our sample who had prior experience with the IV formulation of their current SCOmAb (6/10 patients), all but one (5/6) expressed unprompted that they strongly preferred their current SC treatment, specifically citing benefits such as faster administration times and the convenience of avoiding IV access. The remaining patient did not express any strong preference for the SC versus IV route of administration. However, while all patients in our sample reported that they ultimately tolerate their SCOmAb injections, most (8/10) described that they experience some degree of discomfort during the injection. The types, frequencies, and patient descriptions of discomfort during injections are captured in Table 11, and include discomfort due to needle movement during administration, the total volume of medication injected, and a burning sensation during delivery. Notably, of the patients who did report experiencing discomfort, very few (2/8) noted that they ever voice this to their nurse when it occurs (Fig 2). Of these two instances, one patient requested a change of injection site (to a different location on the thigh) and the other requested an adjustment to the needle angle. In both cases, these patients noted that their nurses responded to their requests and adjusted as a result.

**Fig 2.**
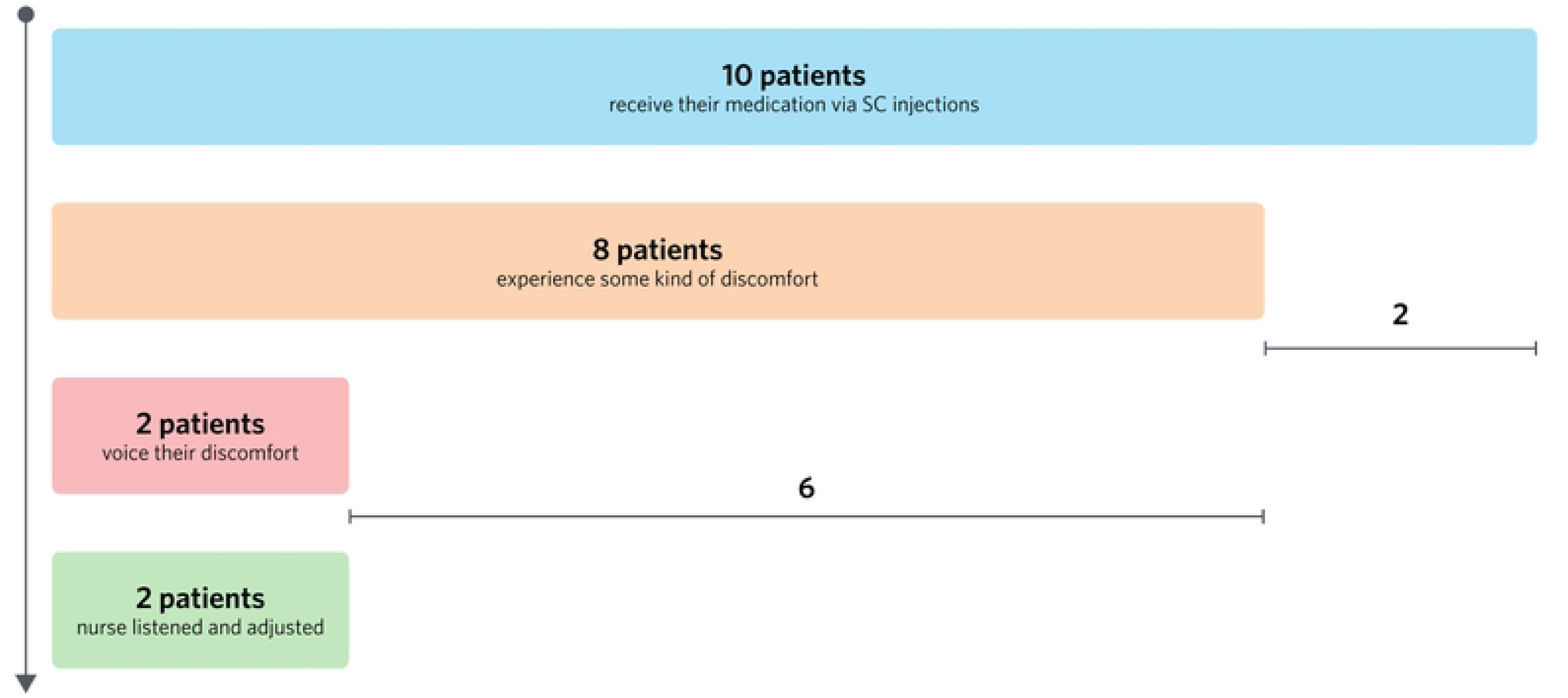
Patient discomfort during injection and communication to nurse or caregiver. (n = 10 patients)

**Table 11.** – Patient Discomfort During Injection (n=8, Patients that experienced any type of discomfort during their injections)

|  |  |  |  |
| --- | --- | --- | --- |
| **Types of Discomfort Experienced by Patients**<br>*(select all that apply)* | **Count for Patients**<br>*(n=8)* | **Quote Identifier** | **Representative Quote** |
| Discomfort from the needle in the skin | 6/8 (75%) | Q14 | “Well I think when I move or if the nurse is not gentle enough. So it’s not exactly the most comfortable thing you’ve ever received because it’s a nice long needle.” – **P9PT** |
| Discomfort from the volume of medication injected | 5/8 (63%) | Q15 | “Some of the people that injected seem to have more experience, and so when they ejected it, it seems like smoother. And then some of the people, when they injected it, seems more like they’re like pushing it harder, or like it’s more jerky I would say.” – **P13PT** |
| Discomfort or burning from the medication itself | 4/8 (50%) | Q16 | “There’s like a, it’s like I can’t put into words. If I could put that sound into words it’d be ‘sssss’ inside.” – **P9PT** |

Nurses shared similar feedback, reporting that they make every effort to provide a positive patient experience and, as noted above, adapt their administration practices for comfort (e.g., use butterfly needles sets, slow/pause injections). As mentioned prior, all nurses reported that if a patient voices pain, they will pause and wait to continue the injection until the patient is ready, but will keep the needle inserted so as not to lose any uninjected medication. In these cases, nurses reported they can usually restart the injection and complete the dose after talking the patient through the discomfort. Other discomfort mitigation strategies described by nurses in our sample included selecting smaller-gauge needles for patients with lower body mass (2/6 reports), using numbing cream or pain medication for patients who are more sensitive to pain (2/6 reports), and taking extra time to answer questions for nervous patients (2/6 reports). All nurses in our sample also reported that they deliberately take time at the beginning of the visit to explain the injection treatment to their patients, answer any questions, and prepare them for the injection.

### Examine how a delivery device would potentially fit current in-clinic workflows

#### Prefilled syringe configurations

Pharmacist/pharmacy technician and nurse participants were presented with two device prospects – replacing the current SCOmAb vial configuration with a PFS, and using a portable infusion pump for SC injections – to assess potential fit of each with existing in-clinic storage, preparation, and administration workflows. Overall, most participants in our sample reported that a PFS configuration would still be stored and distributed by the pharmacy in their facilities (9/10 pharmacist/pharmacy technician reports, and 8/10 nurse reports), although others speculated that having a PFS could trigger a move to automated dispensing cabinets, as similarly packaged medications are already dispensed this way. Many (8/10) of the pharmacists/technicians expected that the preparation process for a PFS would be streamlined and simplified to either attaching an administration needle or dispensing the syringe as is. The remaining (2/10) pharmacist/pharmacy technician participants noted that if dose adjustments are required, the medication may still need to be transferred from the PFS to another syringe (e.g., conventional polypropylene). Many (6/10) nurses agreed that PFS configurations could streamline the dispensing process, citing that they would likely spend less time waiting for pharmacy to prepare medications if they were supplied in PFSs.

#### Portable infusion pumps

As mentioned in a prior section, some nurses in our sample reported that their facilities have already devised methods to automate the SCOmAb injection process using syringe pump modules attached to existing infusion pumps in an attempt to address scheduling and staffing restrictions. One nurse described that her facility uses a makeshift setup that utilizes only materials already stocked in the infusion clinic and repurposes IV supplies for SC use:

> *“We actually have been using our IV tubing. We use an IV set, and we will start the IV [catheter] like subcutaneously kind of with the needle facing up towards the patient [inserted into the patient’s thigh with the catheter pointed proximally towards the patient’s abdomen rather than distally towards the lower leg]. So we’ll clean off the area, insert the needle, put a Tegaderm dressing on it, and then we have just a short tubing that goes up to our syringe pump, which we hook up the Phesgo to and we then program the Phesgo to run over 5 min.” **–*** **P2NR**

Regardless of existing pump use, pharmacist/pharmacy technician and nurse participants were asked about the prospect of using a portable infusion pump for SCOmAb delivery and how such a device would be managed in their facilities. All participants reported that they would expect the portable infusion pump to be stored in the same way infusion pumps are currently stored in their facilities and shared in the same way syringe pump modules are currently shared between patient rooms. Specifically, nurses noted that the pump would be stored in medication rooms or central supply storage rooms, cleaned by the nurse who used it last, and tracked/maintained along with the rest of their pumps by their maintenance/engineering group. In terms of preparation, most (8/10) pharmacists/pharmacy technicians expressed concern that having a portable infusion pump could impact the syringe they would need to use and wondered if a custom syringe or specific size would be needed to fit the pump and avoid errors. Otherwise, they did not foresee any change in their process. Many (6/10) nurses saw potential benefits in using a portable infusion pump to automate the process, noting that it could provide more reliable flowrate control (3/7 reports) and eliminate the physical discomfort they experience during manual syringe administration (3/7 reports). However, incorporating these pumps also raised concerns among all but one (9/10) nurse. Nurses reported that the pump could be cumbersome to use and time-consuming to set up, particularly for smaller dose volumes (5/9 reports), add burden associated with managing the pump itself and stocking necessary components (3/9 reports), require an extra needle securement step (2/9 reports), still require them to stay with the patient while the pump is infusing (2/9 reports), demand additional training on how to use the pump (2/9 reports), and necessitate that reservoirs be filled with more medication to account for residual volume that may be lost in the tubing (1/9 reports). One nurse elaborated on the concern about having to stay with patient during portable infusion pump use, noting that while hands-free administration would offer benefits, she would not feel comfortable leaving the patient unattended during delivery:

> *“I don’t know if I could trust my patient to sit still while that [the portable infusion pump] is going, and I’m away from them. Like I think I would have to be there, but I think that’s great in terms of not having to use your hands to do this. It frees up you as a nurse to, I guess, like talk to the patient.” **–*** **P16NR**

### Project the impact of increased frequency of SCOmAb injections on workload

#### Current time/staffing requirements and future implications

The aggregate treatment timeline described by patients, nurses, and pharmacy staff in our sample is summarized in Fig 3, with relative timing and approximate overlaps between roles shown for comparison. Patients in our sample reported that their total treatment times range from 1.5-6 hours (average 3 hours), inclusive of travel time, wait time, any additional doctor visits or labs, and SCOmAb administration itself. While nurses in our sample reported variability in their injection times compared to SCOmAb prescribing information recommendations as detailed above, the injection itself still represents a small portion of this total time (i.e., on the order of minutes). Of the patients who receive their injections in the clinic, most (6/9) reported that they have labs drawn on the day of their injection, although only some described that their physicians will put holds on administration until lab results are received. Variability with regard to hold parameters was also reported among nurses in our sample, with some portion of the sample reporting that holds are *always* (3/10 reports), *usually* (3/10 reports), *sometimes* (3/10 reports), or *never* (1/10 reports) placed on SCOmAb administration pending lab results.

**Fig 3.**
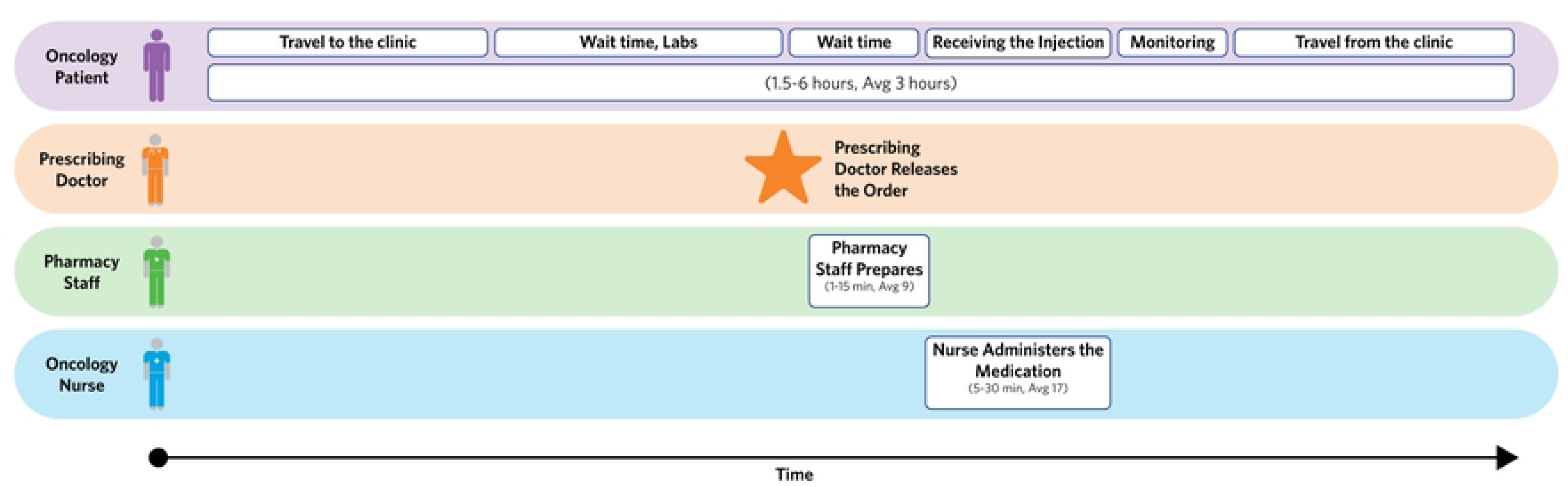
Comparative impact of time and frequency for patients and HCPs.

For pharmacy staff, SCOmAb preparation was reported to be fast and easy compared to more complex IV formulations, typically requiring only 1-15 minutes (average 9 minutes) to prepare a dose. Pharmacists and technicians in our sample described that SCOmAbs are typically prepared for one patient at a time and only after the patient has arrived. Most (9/10) pharmacists/pharmacy technicians reported that their facilities do not batch prepare SCOmAbs, insisting that doing so could result in wasting expensive drug product if patients cancel or cannot receive treatment on their scheduled day (e.g., due to laboratory abnormalities). The one pharmacy staff member in our sample who did report batch preparation only prepares a portion of the SCOmAb doses for that day to ensure all prepared medication is used regardless of any unexpected scheduling changes:

> *“Recently we have [started batch preparing], because the volume has gone up with the patients, so pretty much we’ll do it the night before the last tech, who leaves at about 6 pm, so about 5pm scheduled. A batch of Faspro is maybe up to 10 [preparations], no more than 10.” **–*** **P8PR**

In contrast to pharmacy staff, nurses in our sample reported that SCOmAbs typically require 5-30 minutes (average 17 minutes) to collect supplies and administer a dose, and that the manual nature of the administration requires them to stay with the patient throughout. Half (5/10) of nurses reported this dedicated time creates staffing issues for their facilities and forces them to ask colleagues to cover their other patients during SCOmAb administration. Nurse-reported activities to streamline the SCOmAb administration process are captured in Table 12.

**Table 12.**
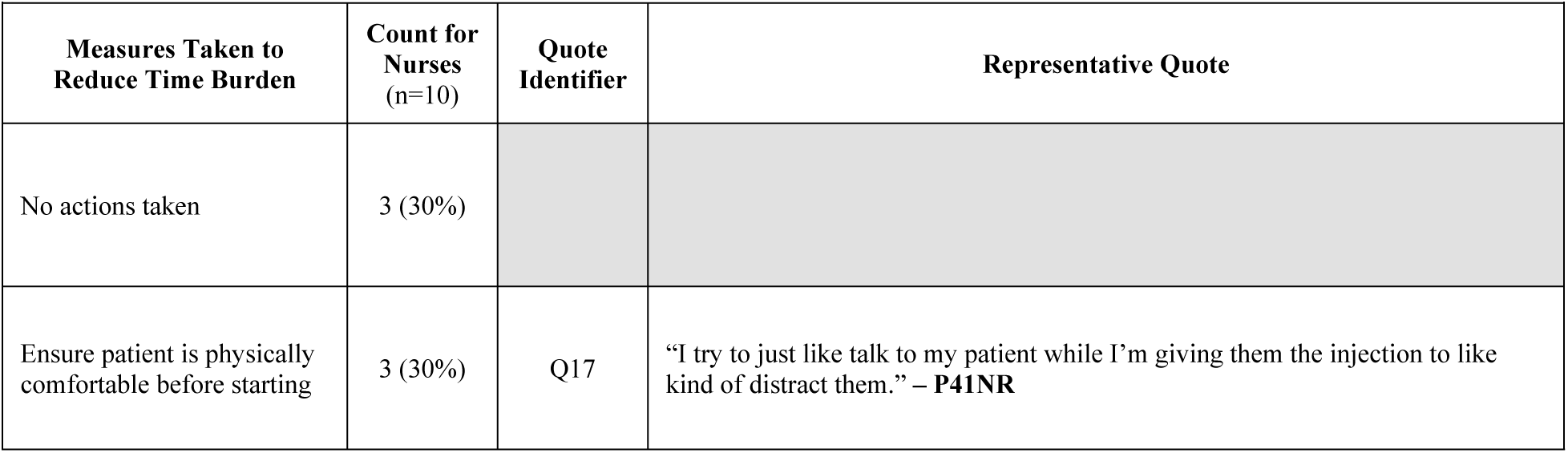
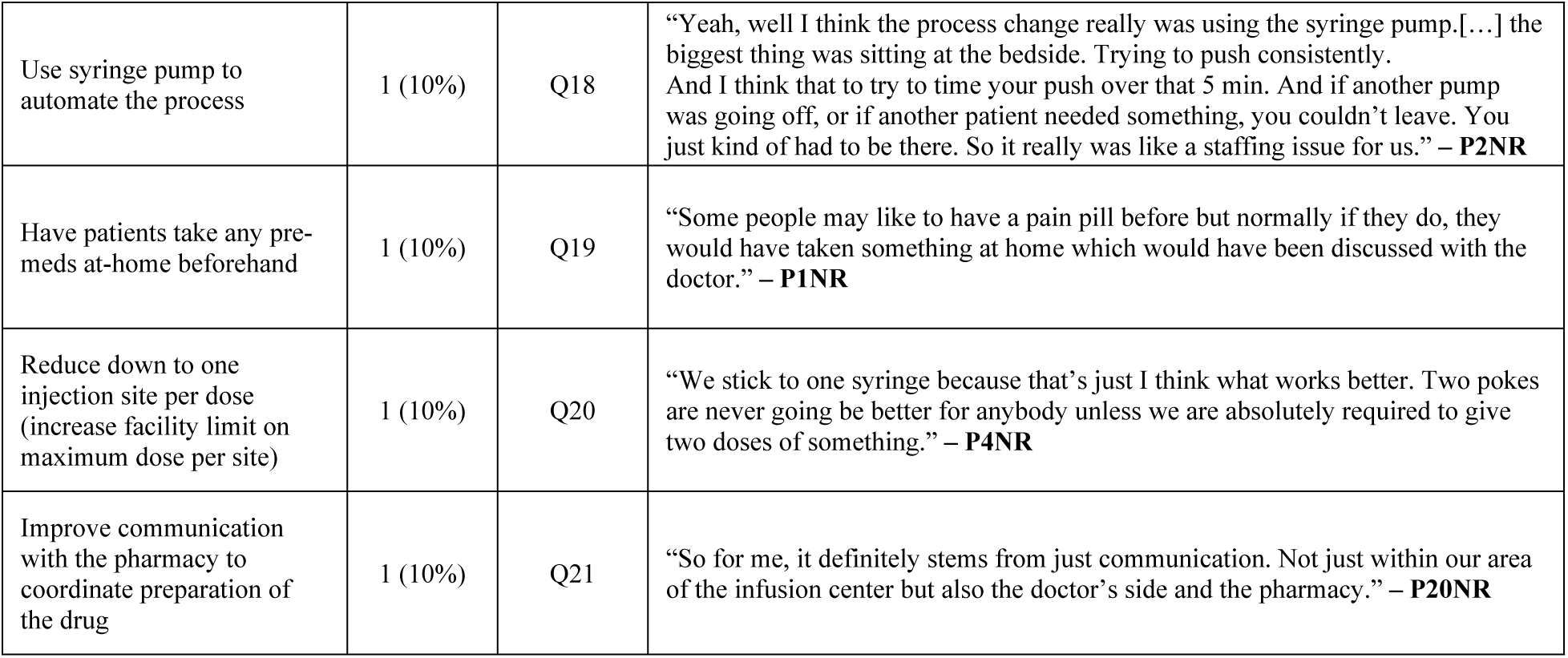
– Measures Taken by Nurses to Streamline Injections and Reduce Time Burden.

Nurses in our sample also emphasized that time and staffing constraints associated with SCOmAbs have been exacerbated by their increasingly prevalent use over time. Some larger clinics (defined in our study as those with more than 50 infusion chairs) reported that individual SCOmAbs can be administered as often as 60 times per day. This rapid adoption was reported to place increased time pressure on nurses and require changes to clinic scheduling and staffing, with most (8/10) nurses stating they would need more staff to cover additional growth. Nurses also projected that they would need added infrastructure to accommodate a greater number of SCOmAb patients, including: more facility space dedicated to SC injections (3/10 reports), additional training for any new SCOmAb agents (3/10 reports), more syringe pump attachments for their IV pumps (2/10 reports), and dedicated pharmacists for SCOmAb preparation to reduce nurse wait times (1/10 report).

### Explore patient and HCP perspectives on home administration of SCOmAbs

#### Current home administration practices

As referenced in Fig 1 and throughout the above sections, one patient in our sample was currently preparing and self-administering a SCOmAb (Herceptin Hylecta) at home at the time of the study. Similarly, one nurse in our sample reported that her facility has begun an initiative to transition roughly one third of SCOmAb patients to home administration. Given that both situations were unexpected recruiting results, likely relatively rare occurrences, and different from the rest of our sample, we focused our interviews with these participants on capturing their unique practices, experiences, and perspectives on home administration. Particular areas of interest included initiation and training, the self-injection process itself (for the patient participant), and other notable differences between at-home and in-clinic administration.

In terms of initiation to self-administration, our patient participant reported that her training was performed with a nurse at the infusion clinic using a syringe and injection pad, where the nurse demonstrated the injection technique. Afterwards, she performed multiple practice injections into the injection pad herself until the training nurse felt comfortable that she understood the process. Once the training process was complete, the patient reported that all of her required materials (medication vials, syringes, and needles) with the exception of alcohol swabs, which she procured herself, were dispensed by her pharmacy:

> *“I just go to the drugstore, hand over my prescription, and they fill it.” –* **P31PT**

Our nurse participant reported that initiation to home administration is facilitated through a conversation between the patient and their physician, who will then order self-administration training once the patient is deemed a viable candidate. The nurse described a similar training process to that reported by our patient participant, explaining that patients will receive training in the clinic and are asked to bring a family member with them to help ensure they retain the necessary information. She went on to detail that patients start by watching a training video and then practice on an injection pad. During this process, the training nurse will provide feedback on the patient’s injection technique, and once they are ready, the patient will perform a real injection on themselves in the clinic before transitioning to home administration.

Regarding the self-injection process itself (only discussed with the patient participant), the patient in our sample described that she sits in her living room to perform the injection and then spends time afterwards lying down to relax. Unlike the typical in-clinic workflow, she reported that she rarely if ever has labs drawn on the injection day, noting that this affords her flexibility and reduces her “treatment day” time to only 10 minutes, as she does not need to travel to a clinic or laboratory facility. The patient emphasized that this flexibility and time savings was the most significant benefit to home administration. Our nurse participant echoed a similar sentiment, citing reduced in-clinic scheduling demands and increased patient convenience as the primary motivations for transitioning patients to the home:

> *“It would be at-home for patients that have difficulty either traveling to the infusion clinic several times a week for a few weeks, or patients that are just wanting to not have to impact their work life by going to an infusion center and investing the time to get the injections.” **–*** **P28NR**

However, despite these benefits, further discussion of the self-injection process with the patient participant revealed latent issues that had the potential to cause preparation and administration challenges. For example, the patient noted that she uses the same needle to draw up (i.e., pierce the vial stopper and withdraw the dose) and administer her medication, which could result in a dulled needle and increased discomfort during the injection. She also reported that the lengthy injection time with manual administration forced her to develop “a stop and go” method to preserve the recommended injection duration without requiring sustained pressure on the syringe plunger:

> *“What I normally do may not be the perfect way to do it. I’ll push for a certain number of seconds, and I count it. Then I stop. Then I’ll start [again]. As long as I don’t pull the needle out, I can’t really do much wrong.” **–*** **P31PT**

The patient went on to describe that she wished the injection was self-regulating to avoid this issue and the need for her adopted technique:

> *“I wish in a way that it had its own governor. […] That it would automatically stop, say, after just a few milligrams, or whatever it is so that you didn’t have to be so careful.” **–*** **P31PT**

#### Benefits and concerns related to home administration

For the remaining patients and HCPs in our sample, the discussion of home administration was inherently projective, as no others had first-hand experience with it. When presented with the prospect of self-administration at home, many (6/9) of the patients in our sample who currently receive their injections in clinic were interested in the potential time savings and flexibility they could be afforded. Many (5/9) specifically mentioned that avoiding transportation to and from the clinic would be a benefit and others (2/9) reported they would prefer the more relaxed environment at home. Still, most (8/9) also expressed concerns about self-administration at home with the current SCOmAb presentations, including: uncertainty about what to do if something went wrong with the injection (4/8 reports), fear of self-administering with the syringe (3/8 reports), apprehension about storing the medication at home (1/8 reports), and difficulty ensuring sterility during self-injection (1/8 report). Patients emphasized that dedicated training and support would be required to navigate these potential challenges.

Nurses in our sample shared comparable concerns when presented with the idea of patient self-administration, especially considering the current manual push administration process. All nurses expected that their patients would face some challenges, specifically: knowing who to contact with issues (2/10 reports), not having anyone monitoring for side effects or adverse reactions (3/10 reports), not having the strength to administer via manual push (3/10 reports), not having the appropriate knowledge for self-administration (2/10 reports), or having a language barrier that impacts training (1/10 reports). Similar to patient sentiments, all nurses also stressed that a thorough training process would be required with current SCOmAb presentations, either through in-clinic sessions (6/10 reports) or at-home sessions (4/10 reports). The importance of thorough training, appropriate patient selection, and close follow-up was echoed by the nurse participant whose facility often transitions patients to self-injection at home:

> *“So in addition to the initial training, I think it’s always important for like a follow up. Like I used to call people the next day when they were gonna do their first [injection] and say hey are you good, like do you have questions? […] I don’t think these types of things are for everybody, and I think there would be some patients that would struggle. We have seen those patients, so our current programs for anybody going home with anything is that they have to demonstrate the ability to do so.” **–*** **P4NR**

## Discussion

This study examined how SCOmAbs have been integrated into the workflows of outpatient oncology infusion centers by capturing the perspectives of pharmacists/pharmacy technicians, nurses, and patients across diverse facilities. The inclusion of all three stakeholder groups was deliberate, as each interacts with a different segment of the SCOmAb workflow and therefore offers a distinct vantage point on where current processes work well, where challenges exist, and where improvements could have the greatest impact.

Several key findings emerged from this analysis, each with implications for how current SCOmAb products are used in practice and how future product presentations and delivery platforms might be designed to better support clinical workflows. While SC administration clearly reduces administration times compared to IV delivery, our findings suggest that meaningful opportunities remain for further streamlining and simplification when the entirety of the preparation, administration, and care coordination process is considered. These findings are summarized below and discussed in the subsequent sections.

1. **Preparation practices are generally well-integrated into pharmacy workflows but vary considerably across facilities, particularly with regard to use of supplies and location of preparation.** These findings suggest that future product presentations should accommodate diverse preparation workflows, including both centralized pharmacy compounding and emerging bedside preparation models, while ensuring compatibility with CSTDs where facility policy requires their use.
2. **Nurses are adapting administration supplies and techniques beyond what is described in product labeling, including use of non-standard needle types, syringe pump attachments, and IV catheters for SC delivery.** These facility-level workarounds indicate that current manual syringe presentations do not optimally match clinical workflow needs for a subset of users, and that purpose-built delivery solutions designed specifically for LVSC administration could address the underlying challenges that have prompted these adaptations.
3. **Manual push administration creates ergonomic challenges for nurses and limits their ability to multitask during delivery, with both issues expected to intensify as SCOmAb utilization increases.** The combination of sustained physical effort, restricted positioning, and prolonged “tethering” to patients during administration points to a clear opportunity for improvement. Delivery mechanisms capable of reducing or eliminating manual effort, while restoring the hands-free time that characterizes IV pump-based workflows, could meaningfully address both the ergonomic and operational constraints identified in this study.
4. **SCOmAb injection rates in practice are driven more by patient tolerance and nurse judgment than by product labeling alone, with nurses in our sample consistently adjusting pace and pausing delivery based on individual patient response rather than adhering strictly to labeled administration times.** These findings suggest that any future automated delivery methods for SCOmAbs should preserve the ability to responsively adjust or pause delivery without requiring sustained physical effort. In particular, passive or instant-response pause mechanisms may be preferable to those requiring active user input to maintain cessation of flow. More broadly, the degree of rate control afforded to clinicians and patients should be a deliberate design consideration for any delivery device intended for SCOmAb administration, as the flexibility that characterizes current manual practice may not be easily replicated by devices with fixed or narrowly defined delivery profiles.
5. **Both patients and nurses are interested in home self-administration of SCOmAbs for time savings and convenience, but also raise significant concerns about performing the current manual syringe injection process independently.** This was not purely hypothetical in our study, as participants with direct home administration experience confirmed both the feasibility of home transition and the practical challenges imposed by the current manual syringe process, including the need for extensive individualized training and patient-developed workarounds to manage the physical demands of injection. Of course, patients outside of oncology routinely self-administer SC therapies at home with injection devices, often despite suboptimal training, suggesting that the barrier to broader home adoption of SCOmAbs may be rooted more in the current product presentation than in the concept of home administration itself. Purpose-built delivery devices that prioritize intuitive operation, minimize the number of user steps, and reduce the physical demands of administration could therefore lower both the training burden and the threshold for safe home transition.
6. **Nurses anticipate that further adoption of SCOmAbs will increase strain on infusion center operations and necessitate additional staffing, training, and infrastructure, and early indications from pharmacy practice suggest that preparation workflows may also be affected as volumes grow.** As SCOmAb utilization expands, including the potential for multi-drug SC regimens, delivery platforms that reduce per-administration resource burden and accommodate administration of multiple medications could help maintain nursing throughput and scalability. On the pharmacy side, growing volumes may increasingly push facilities toward batch preparation practices, raising concerns about cost of waste, in-syringe stability, and prolonged drug contact with unintended materials that prefilled product presentations could alleviate entirely.

### Current practices with SCOmAbs delivered in clinic

#### Preparation

Overall, participants described that SCOmAbs are usually prepared in typical pharmacy environments (i.e., compounding suites) and follow standard pharmacy preparation procedures. Pharmacy staff particularly emphasized that these preparations are less complex and time-consuming compared to many IV preparations. This observation is consistent with a systematic review of 72 studies comparing time and resource differences between SC- and IV-administered oncology biologics, which concluded that SC formulations save between 5.6 and 21 minutes of pharmacist/preparation time.[8] A recent real-world evaluation of Phesgo implementation in Japan further corroborated these time savings, demonstrating reductions in active treatment time of up to 96% and cumulative savings of over 330 hours of chemotherapy suite time across 8 months of use, with nurses and pharmacists reporting reduced task complexity and improved workflow satisfaction.[77] Other more qualitative accounts also support the relative ease of preparation of SCOmAbs versus their IV alternatives, citing benefits such as avoidance of dose calculation, dissolution, and dilution steps.[78,79]

The well-established and largely trouble-free SCOmAb preparation process in pharmacy made nurse bedside preparation a somewhat surprising finding in our study, especially considering preparation of most oncology therapies is specifically managed by pharmacy due to their criticality and risk of toxicity.[80–83] There are of course also existing models for bedside preparation in the oncology setting, although these are often centered on supportive treatments (e.g., antiemetics, corticosteroids, antihistamines, antipyretics, analgesics). In our study, it appears that given their comparable preparation requirements, some facilities treated SCOmAbs similarly to these more commonly nurse-prepared medications that can be safely and reliably managed at the point of care after dispensing via automated dispensing cabinets. The concept of storing SCOmAbs in clinical areas rather than in pharmacies has also been suggested elsewhere as a means to reduce burden on pharmacy compounding and eliminate wait times for treatments to arrive at the bedside.[84] Although not observed in our sample, another study specifically evaluated nurse-led preparation of oncology mAbs, including IV formulations, in clinical areas using CSTDs to prevent needlestick injuries, spillage or aerosolization, and contamination during preparation.[85] Of note, this initiative was designed to offset increasing demand for oncology biologics and the limited capacity of the affiliated pharmacy to manage additional preparations. Still, transitioning preparation to the bedside could also increase workload for nurses and/or require dedicated space that may not be readily available on the unit as knock-on effects,[84,86] and the UK Systemic Anti-Cancer Therapy (SACT) Board recommends that this practice should be explored cautiously and after other options to reduce demand on pharmacy preparation have been properly explored[87] to avoid simply shifting pressure from one practice area to another. Future studies should specifically seek to understand the criteria for decentralizing storage (i.e., in automated dispensing cabinets) and preparation of SCOmAbs, as doing so elegantly has the potential to streamline both pharmacy and nursing workflows and therefore reduce patient time in clinic. Moreover, the development of fully prefilled device solutions (e.g., PFS; discussed in more detail later) could minimize the concern about shifting preparation burden by removing preparation steps entirely.

Use of CSTDs for SCOmAbs preparation and overall treatment of these products as hazardous drugs were also topics of interest in our study, given the recent drug compatibility concerns raised by the pharmaceutical industry working groups.[34–36] Even among our small sample, we observed significant variability in CSTD utilization and several participants reported issues associated with their use, such as potential for these components to break, introduce air bubbles into the preparation, or create more unwieldy systems due to added length with adaptors. Interestingly, we also observed some disconnect in our sample between the use of CSTDs and other precautions intended to prevent hazardous drug exposure, suggesting lack of strong consensus on whether SCOmAbs are truly considered hazardous in these facilities. This was not altogether surprising, as other authors have indicated that although most oncology biologics are not considered hazardous per the NIOSH List, they are still sometimes treated as such.[32,33] Another logical explanation is that facilities abandon hazardous drug precautions for SCOmAbs at different steps in the workflow (e.g., administration), realizing that it is not technically possible to ensure a completely closed system during the full use process when a needle must ultimately be connected for SC delivery. This preconception could result in increased exposure risk if products are prepared at bedside, however, and the UK SACT Board specifically recommends using CSTDs when preparing mAbs outside an aseptic unit.[87]

Regardless, pharmaceutical manufactures should continue to recognize that CSTDs may still be used (albeit inconsistently) with their current and future product presentations, and that compatibility with this workflow must be minimally assured for adoption. This may be particularly relevant if new delivery device configurations that require transferring from current vial presentations are introduced, as such systems would need to be compatible with on-market CSTDs or ensure aerosol-free transfer can be maintained by the device itself.

#### Transport

Following preparation, participants described how the finished product is closed and transported to the point of care as a segue to administration practices. Here, the most commonly reported process was for pharmacy to prepare SCOmAbs to be as “ready-to-use” for nurses as possible, which even included attaching an administration needle in some cases. This finding is consistent with overall pharmacy compounding best practices, where ready-to-administer products prepared under the supervision of pharmacists are considered vital to improving patient safety.[88] However, it is worth noting that all currently approved SCOmAb prescribing information instructs that the hypodermic injection needle or SC administration set should be attached to the syringe immediately prior to administration to avoid potential needle clogging, which would make pre-attachment during pharmacy preparation technically inconsistent with labeled instructions. While this practice may reflect a pragmatic effort to streamline workflow and was not reported to cause issues in our sample, pharmaceutical manufacturers should be aware that it occurs and consider whether future product designs or labeling could better accommodate this observed workflow pattern. For others in our sample, administration supplies were designated to the nurse, which can be likened to nurse “ownership” of the administration process for IV infusions, including selection and connection of IV tubing, catheter management, and infusion pump programming. In terms of transport to the bedside itself, practices were highly facility-specific and participants reported that a variety of methods may be used, as other authors have also described for mAbs.[89,90] Two techniques employed in our study that may be of particular interest are the use of pneumatic tube systems and dumbwaiters, as both impose size constraints and may introduce mechanical agitation that could potentially impact mAb stability.[91] Like CSTD use, pharmaceutical manufacturers need to be aware that their medications will likely be subjected to variable transport methods, and product configurations should ideally be intentionally designed with these in mind.

#### Administration

SCOmAb administration practices in our sample were largely dictated by facility protocols, with some aspects dependent on nurse preference. Notably, no nurse participant made particular mention of product labeling requirements when discussing administration, although it is likely that these were already reflected during development of facility protocols. Instead, nurses described that administration practices centered on practicality, with workarounds developed to prevent errors (e.g., using larger syringes than necessary), anticipate challenges (e.g., proactively splitting doses), or troubleshoot situationally, even if they are not “correct” per product labeling or best practice consensus. This disconnect between “correct” and “practical” has been discussed extensively in the literature.[39–42]

Adaptations in our study were specifically noted with regard to administration supply selection. Most nurses in our sample described using supply variants (i.e., syringe sizes, needle types, needle gauges) that are consistent with prescribing information (Table 1) although some reported using entirely different supplies and techniques not described in product labeling (i.e., use of IV catheters and syringe pumps). While there is limited literature specific to supply selection for administration of SCOmAbs, one mixed-methods study of SC versus IV trastuzumab administration found that the ability to choose smaller needles based on patient preference was a positive feature of the SC formulation.[79] Conversely, nurses in our sample expressed that they often select the *largest* needle that patients can tolerate in order to reduce the injection force and speed up the administration. Regardless, the use of larger needles, butterfly needles, and alternative methods like IV catheters and syringe pumps in our study and elsewhere[43–45] serve as an initial indication that the current manual administration method can be difficult to perform, at least for a subset of nurses. Additional pain points related to this process as reported in our study are discussed in the next section.

### Identified pain points with current SCOmAb administration

#### Injection time

Nurse feedback about their chosen injection rates and timekeeping methods revealed an overall lack of precision associated with the SCOmAb administration process, without necessarily any regard for product labeling recommendations. Although not specifically stated by nurses in our sample, this may be related at least in part to the use of ranges or approximations to describe injection times in the labeling itself (see Table 1), which reflects that some nominal degree of variation is acceptable. A recent report comparing SC versus IV administration of atezolizumab confirmed this behavior, as injection durations for SC atezolizumab were found to range from 2 minutes to 14 minutes (median 7.1 minutes and average 4-8 minutes for 75.7% of patients).[92] Of course, it must be noted that this variation did not impact pharmacokinetics, safety, or efficacy, nor would any impact be expected when administration rates are varied for most SC biotherapeutics.[93–98]

Injection time is inherently flexible in SCOmAb labeling, and nurses in our study unsurprisingly treated it as a variable they could control. This is reasonable given that SCOmAb labels describe injection time in terms of an approximate *duration* (e.g., “over approximately 8 minutes”) rather than a precise *rate* (e.g., “at 1 mL/min”). This is not particularly unusual, as administration durations are commonly used for a variety of IV medications as an alternative to precise infusion rates. However, in these cases, rates can still be easily calculated and controlled with the help of infusion pumps with minimal user discretion, which is not the case for manual syringe administration with SCOmAbs. Moreover, nurses in our sample expressed that they sometimes actively deviate from SCOmAb prescribing information recommendations based on patient tolerability or facility protocols. This reinforces that patient-specific factors can meaningfully affect SC injection tolerability, and there is a general lack of consensus on “standard,” “acceptable,” or “ideal” SC administration rates, particularly for large molecules.[99] All nurses in our sample reported that they will pause the injection in the event of patient discomfort or other adverse event and wait for the patient to be ready to continue before resuming. This practice is consistent with prescribing information recommendations for some SCOmAbs.[25–27,30] and reports from clinical practice where decreasing the dosing rate was specified as a coping strategy for patients experiencing injection site reactions during SCOmAb administration.[100] While this is possible with manual syringe administration, it is not for many of the automated SC injection devices (e.g., autoinjectors [AIs], OBDS) currently used outside the clinic, suggesting that any future automated delivery methods for SCOmAbs should consider the potential benefit of being able to pause delivery when needed. This function should be passive for the administering nurse, however, as the need to press and continue to hold a button to pause an injection (“active pause”) has been specifically raised as a potential limitation of an OBDS under investigation for SCOmAb delivery.[101]

#### Injection ergonomics

Nurse positioning during the injection was largely described as a learned behavior in our study, with most nurses reporting that they’ve adopted a comfortable sitting position over time after finding more typical standing positions uncomfortable. Moreover, nurses expected their own and their patients’ positioning to remain consistent throughout the injection, even when butterfly needles were used to “decouple” the injection site from the syringe and ostensibly allow for more flexibility. The injection technique itself (i.e., continuous versus pulsatile injection) was similarly reported to be an adopted behavior, this time mostly according to practices with other medications, as the recommended injection technique for SCOmAbs is notably not specified in product labeling. Continuous injection was reported most commonly, which is consistent with delivery profiles of automated SC injection devices used outside the clinic (e.g., AIs, OBDSs), although these devices certainly offer more consistent delivery profiles compared to manual administration.[102–104]

Regardless of physical positioning or injection technique, the majority of nurses reported that they experience some degree of physical discomfort or pain associated with the injection process. Although the presence or absence of MSDs could not be determined through this qualitative study, nor was it the intent, the annual prevalence of MSDs among the nursing profession has been found to be considerably high (77.2%), with the lower back (59.5%), neck (53.0%), and shoulder (46.8%) the most affected anatomical areas.[50] Similar concerns have been raised with regard to repetitive and continuous motions associated with syringe use for manual administration of IV chemotherapy[47] and SC trastuzumab.[78] While not all of these findings are specifically related to SCOmAb administration, it is possible that the complaints raised by nurses in our study could signal similar ergonomic challenges and risk of occupational injuries. At a minimum, the need for specific positioning, restricted movement, “tethering” of the nurse to the patient during manual delivery, and corresponding complaints of pain or discomfort suggest that current SCOmAb administration represents a more physically challenging procedure compared to other medications typically delivered in clinic. Future products that can be delivered more comfortably, more quickly, or entirely hands-free could therefore have the potential to improve upon the ergonomic challenges raised in this study.

#### Impact of large volumes and high viscosities

High viscosity is a common and well-known characteristic of biologic formulations that can result in administration challenges for affected products. As many biologic molecules exhibit an exponential relationship between concentration and viscosity, increasing formulation concentrations to shrink dose volumes can produce products with very high viscosities.[20] Moreover, SCOmAb volumes are already notably higher than those of most SC-administered biologics. Due to both of these factors, we hypothesized that the combination of higher viscosities compared to water-like drugs and higher volumes compared to most biologics could present potential challenges for clinical staff who prepare and administer SCOmAbs.

This was not the case for pharmacists/pharmacy technicians, who did not consider 5-15 mL dose volumes to be particularly large compared to other products they routinely prepare. Similarly, they also reported that they do not perceive SCOmAbs to be particularly viscous, commenting that the ability to use large needles in preparation makes manipulating these medications less challenging. Increased perceived viscosity of the SC fixed-dose combination of pertuzumab and trastuzumab compared to its IV alternatives has been described in one study, but augmenting supplies was similarly suggested as a means to minimize the potential impact.[22] In contrast, nurses in our study collectively expressed a different sentiment to pharmacy staff, with most noting that large volumes and high viscosities are contributors to overall SCOmAb administration difficulty.

In an effort to contextualize nurse feedback, the viscosity of several on-market SCOmAbs and their associated glide forces (injection forces) in a syringe were measured and calculated as a supplement to the core study results (S1 Table). Notably, the SCOmAbs measured have reasonably moderate viscosities (4-7 cP) and formulation concentrations (all are also co-formulated with hyaluronidase, which is not expected to increase viscosity), requiring a user to push with an average force of 7 N (4-10 N range) for the syringes used in our test lab. These injection forces are in a similar range to what has been observed with smaller-volume, on-market biotherapeutics;[105] however, the injection times are considerably longer due to their larger volumes, which may have contributed to the feedback related to hand strain and overall administration difficulty in our study. With this in mind, developers of larger volume SCOmAb products should consider both glide force and endurance when evaluating product usability and the ability of users to deliver the product.

#### Patient experience

As mentioned above, the goal of this study was not to specifically compare patient experience with SC vs. IV oncology mAbs, as this has already been described in detail by prior research. With that said, patients in our sample who had experience with IV mAbs volunteered that they overwhelmingly preferred the SC route for reasons consistent with prior literature.[1,15,106] Despite their prevailing preference, patients also noted discomfort with SC administration related to needle movement during administration, the total volume of medication injected, and burning sensations from the medications themselves.

Unsurprisingly, nurses in our sample heavily prioritized providing a positive treatment experience for their patients on injection day, including adjusting their usual workflows to accommodate individual patient needs or adapting their administration practices for comfort. Although we did not enroll nurse-patient dyads, the patients in our sample independently confirmed that nurse behavior can indeed affect their treatment experience either positively or negatively. Future dyadic research may be warranted to further explore the impact of specific nurse behavior on their patients’ experience during SCOmAb treatment.

### Delivery device fit

#### Replacing vials with PFS configurations

Pharmacists/pharmacy technicians and nurses responded very positively overall to the notion of replacing current SCOmAb vial presentations with PFS presentations. This was primarily driven by the expectation that a PFS configuration would streamline both the preparation and administration processes, with positive impact on the complete workflow. Given the widespread use of prefilled products (e.g., antibiotics, vasopressors, anticoagulants, analgesics) in clinical settings already, it is likely this assessment was rooted in first-hand experience rather than speculation, as existing literature does support the potential efficiency, safety, and drug waste benefits of prefilled products.[107] Although not mentioned by participants, replacing current vials with PFS configurations may also offer benefits to those facilities who use CSTDs to prepare and/or administer SCOmAbs, as fewer required manipulations could streamline the process and reduce opportunities for drug exposure. This benefit has also been described in a UK-based initiative focused on decentralizing delivery of SC atezolizumab, where prefilled syringes were acquired from an external compounder, therefore precluding preparation from vials and associated exposure risk.[84] Lastly, a PFS presentation, especially of a smaller dose volume, may also facilitate delivery in less formal or intensive environments (e.g., a clinical exam room vs. infusion room) and without the need for specially trained staff,[108] potentially freeing up needed chairs for more complex administrations. This could be particularly beneficial for facilities who are already allocating designated areas solely for SC administration to maximize throughput.[84]

#### Automating administration with portable infusion pumps

The idea of adopting a portable infusion pump for SCOmAb administration raised both potential benefits and concerns among pharmacist/technician and nurse participants. Pharmacists/technicians were generally neutral on the concept, as most of the potential influence of a portable infusion pump would occur outside of their purview (i.e., during administration), with minimal impact on their overall preparation steps. With that said, they did express a consistent concern centered around the need for specific syringes to ensure pump compatibility and the potential for malfunctions if existing or similar-appearing syringes were used by mistake. While this may be more of a challenge with pumps that require fully custom syringes compared to others that do not,[109] it is nevertheless a reasonable concern, as pump errors resulting from incorrect component selection have been reported in the literature.[110] Overall, the implication of pharmacist/pharmacy technician feedback is that introduction of any device should not introduce radically new or more challenging preparation steps into their workflow. While they are now used in the treatment of cancer and other therapeutic areas, elastomeric pumps may represent analogous examples, as these devices require considerable force to fill and are challenging and time-consuming to prepare.[111,112]

Unlike pharmacists/technicians, nurses in our sample foresaw several potential benefits of automating SCOmAb delivery with a portable infusion pump. These were primarily related to addressing the limitations of the current manual administration process, and included the potential to standardize delivery rates with the device and ease their own physical burden during administration. Similar proposed benefits have been reflected in a study by Ammor et al., where a syringe pump configuration incorporating a three-way stopcock and IV/SC catheter was scored by nurses as the most acceptable method for SC daratumumab administration compared to other alternatives, although the exact rationale for this preference was not described.[43] Of interest, several nurses in our study also viewed reliable rate control as a prerequisite for adoption of any pump for SC administration, insisting that it should provide a consistent flowrate accuracy to ensure the overall delivery time is predictable. While not specified by nurses directly, this preference may have been related to the overarching desire to systematize the workflow and time allocations for each patient for scheduling and staffing purposes, which was a persistently described theme throughout our study. Another hypothesis could be related to negative experiences with devices such as elastomeric or other mechanical pumps, which are known to have inconsistent flowrates in different environmental conditions.[113,114] Indeed, a recent multicenter survey of 33 nurses across nine Danish hospitals using a non-electronic, spring-driven mechanical infusion system for short-duration SC infusions reported that most experienced less hand strain (81%) and had more time for patient interaction (97%) compared to manual push,[115] broadly consistent with the benefits projected by nurses in our study, although the specific therapeutic context was not described. However, the same survey also reported that 24% of nurses experienced at least one device malfunction and an additional 12% observed temporary slowdowns in infusion attributed to the device’s adaptive flow mechanism, which automatically reduces flow in response to increased back-pressure at the infusion site. While the authors characterized this latter behavior as an intended safety feature rather than a malfunction, it nonetheless illustrates that even purpose-built mechanical pumps may exhibit the type of flowrate variability that nurses in our study specifically cited as a prerequisite concern.

Along with the benefits of infusion pumps, nurses also raised potential concerns about adoption. These ranged from increased time and burden associated with managing the pump and its required supplies to requirements for additional steps, such as training, needle securement, and reservoir overfilling. Some nurses also felt that they would still need to remain at the bedside with the patient even if a pump was used, concerned that patients could inadvertently dislodge the needle during unsupervised delivery, therefore detracting from the efficiency benefit that using a pump could provide. Concerns related to pumps being cumbersome and requiring additional storage requirements, components, or supplies suggest that any delivery device must fit seamlessly with existing pharmacy and nursing workflows to be compatible with infusion center operations. Although not specific to SC administration, one study identified that inadequate availability of required supplies, disconnects in responsibility for supplying, cleaning, and maintaining items, and ambiguity about whether supplies of necessary equipment were sufficient were major contributors to operational failures in a hospital setting.[116] Further, the same study found that these failures often forced nurses to compensate with workarounds, such as “going shopping” in utility rooms to find surrogate equipment. This practice could entirely compromise the function of delivery devices that rely on careful selection of components and potentially lead to administration errors, as have been reported with elastomeric infusion pumps.[117] If infusion pumps are to be more widely adopted for SCOmAb delivery, future research is warranted to establish the ideal pump characteristics for this application to maximize the benefits, alleviate the concerns raised by clinical staff in our study, and prevent known issues associated with drug delivery devices used in clinical settings. As with other delivery devices discussed later in this section, the value proposition of a portable infusion pump is also likely dependent on the specific SCOmAb being administered, as products with shorter injection times and lower manual delivery forces may not present sufficient administration burden to justify the added complexity of pump setup and management. KORU Medical Systems recently submitted a 510(k) premarket notification seeking FDA clearance for use of their FreedomEDGE^®^ mechanical infusion system to subcutaneously administer Phesgo,[64] which would represent the first regulatory-cleared portable infusion pump indicated for delivery of a SCOmAb if approved. In a related finding, a recent retrospective study comparing infusion pump-assisted versus traditional manual nurse push for Phesgo administration found that pump-assisted delivery was associated with significantly lower rates of postinjection pain, swelling, induration, and SC bleeding compared to manual injection.[118] Although preliminary, these results suggest that the controlled, constant-rate delivery enabled by pump-assisted injection may offer tangible patient comfort benefits over manual push, further supporting the rationale for investigating standardized delivery mechanisms for SCOmAb administration.

#### Administration with on-body delivery systems

OBDSs represent another device class that could potentially address the ergonomic and workflow challenges identified in this study. Given the considerable recent attention OBDSs have received as potential solutions for LVSC administration, the emerging evidence base for these devices in the SCOmAb context warrants more detailed examination. OBDSs are generally designed to be affixed to the patient’s body and deliver medication subcutaneously without requiring continuous manual effort from the user, thereby offering hands-free administration that could alleviate tethering, reduce physical strain, and free nurses to attend to other patients during delivery. However, OBDS designs vary considerably in their mechanisms (e.g., spring-driven, elastomeric, electromechanical), filling configurations (e.g., user-filled from vials, prefilled by the manufacturer), and complexity, each of which carries distinct implications for preparation workflows, training requirements, and overall fit with clinical operations. As such, the potential benefits of any individual OBDS must be weighed against the practical demands it introduces across the full use process. To date, the most substantial published evidence for OBDS use with SCOmAbs comes from clinical investigations with a single device platform, which is discussed below.

Since the completion of this study, several publications have described experiences with an investigational, single-use, disposable, elastomeric, user-filled OBDS for administration of SC isatuximab,[66,67,101], which has since been recommended for EU approval,[119] providing an additional lens through which to contextualize the device-related perspectives captured here. Based on the workflow described in one of these studies (Avello et al.), the OBDS was presumed to be filled by pharmacy staff in clinical trials using conventional syringes and then transported to the administering nurse, although other filling options have also been alluded to.[120] While the studied OBDS was preferred to manual administration by HCPs[101,121] and patients[121] and offered the benefit of hands-free administration, addressing some of the strength, ergonomic, and tethering challenges identified by nurses in our study, a device configuration that requires user filling may also introduce preparation steps that could be burdensome[111,112] or similarly physically demanding,[47,122,123] particularly if they are new, different, or more challenging to perform compared to standard practice. This is an important consideration in light of our own findings and those from other initiatives centered on SCOmAb decentralization,[84,86] where the notion of transitioning preparation from pharmacy to the bedside raised concerns about shifting workload between practice areas.

Separately, two surveys sponsored by the device manufacturer assessed pharmacist and nurse preferences for the OBDS compared to conventional syringes for LVSC drug preparation and administration.[124,125] In both surveys, the majority of respondents reported a preference for the OBDS, with pharmacists endorsing potential improvements in efficiency, burden reduction, and preparation safety,[124] and nurses surmising that the OBDS could improve clinic throughput better than syringes[125] when presented with hypothetical scenarios. However, both surveys relied on written descriptions of a device identified only as “Product X” rather than direct device handling, and methodological concerns including hypothetical bias, sampling approach, and omission of practical workflow parameters have been raised by other authors.[126] In contrast, a survey of 12 clinical trial nurses with direct experience administering SC isatuximab via the OBDS reported more measured assessments of the device’s incremental value over manual syringe delivery.[101] While all nurses agreed the OBDS improved efficiency and was easy to learn, perceived benefits relative to SC manual push specifically were modest: 60% agreed the OBDS could reduce time patients spend in clinic compared to manual SC delivery, 50% agreed it could improve patient comfort, and 50% agreed it could decrease patient anxiety.[101] Moreover, nurses with hands-on experience identified practical concerns in free-text responses, including the size and bulk of the device, the need for dedicated training and an associated learning curve, lack of trust in the automation of SC delivery, risk of allergy to the adhesive tape, and positional challenges for mobility-limited patients. The majority (83%) also agreed that uncertainty about the amount of drug delivered if an injection is interrupted could represent a device limitation,[101] which may echo the feedback from nurses in our study who reported that controlled pausing is a necessity in the event of injection pain.

Taken together, these data suggest that while OBDSs may address specific pain points associated with manual SCOmAb administration, they may also introduce their own workflow challenges similar to those described throughout this study. The incremental benefits reported by nurses with hands-on OBDS experience in these studies, combined with the practical concerns they raised, are broadly consistent with the mixed reception of portable infusion pumps described by participants in our study and reinforce that any device introduced into the SCOmAb workflow must be evaluated in terms of its net impact across the full preparation and administration continuum rather than administration benefits alone. Moreover, the value proposition of hands-free administration is likely not uniform across all SCOmAbs, as products with shorter injection times and lower volumes (e.g., trastuzumab at 5 mL over 2 to 5 minutes, or pembrolizumab at up to 4.8 mL over approximately 2 minutes) may not present the same degree of administration burden that would justify the added complexity of a delivery device. In contrast, products requiring larger volumes administered over longer durations may represent use cases where the ergonomic, workflow, and tethering challenges identified in this study are most pronounced and where the tradeoffs associated with device adoption are more clearly favorable. With that said, the exact threshold at which the benefits of device-mediated delivery sufficiently outweigh the added complexity of device adoption is not yet understood and warrants further investigation, ideally informed by the perspectives of the HCPs and patients who navigate these tradeoffs in practice.

### Projection of increased SCOmAb frequency

Although this study was not designed to directly compare IV and SC workflows, the preparation, administration, and overall treatment times reported by participants were largely in line with systematic reviews comparing the time and resource requirements of IV versus SC oncology biologics.[8,15] This consistency with prior evidence reinforces the already well-established operational benefits of SCOmAbs. With those benefits as context, our interviews instead focused on aspects of the SCOmAb workflow that have not been well explored in prior studies, including how preparation and administration processes themselves impact in-clinic workflows and what consequences could result from increased SCOmAb adoption over time. Notably, the same time savings that make SCOmAbs attractive may also contribute to higher patient throughput, which, while clearly a benefit of SC administration, can simultaneously amplify the per-administration workflow and staffing challenges identified in this study.

#### Current and future impact on pharmacy staff

As described above, pharmacy staff reported that SCOmAb preparation requirements have minimal impact on their workflows since these products are generally simpler and take less time compared to other oncology preparations. However, one area of pharmacy practice that could be impacted by increased SCOmAb utilization is the propensity to batch prepare these products. Batch preparation, also referred to as pre-preparation, premixing, or advanced preparation, has been successfully employed as a strategy to reduce oncology pharmacy turnaround times, although medications with short stability and/or high-cost, including mAbs, have been typically excluded.[81,82,127,128] This was true in our sample as well, with all but one pharmacy staff member refraining from batch preparation due to concerns about the cost of waste. However, the one pharmacy staff member who did perform batch preparation in our sample reported that this change was necessary to accommodate high volumes of SCOmAb patients, with minimal risk that pre-prepared products would go to waste. Given the significant pipeline activity in SC oncology,[56] it is possible that premixing could become a more standard practice to meet future preparation demands. While currently FDA-approved SCOmAbs can likely accommodate this based on their labeled in-syringe stabilities (see Table 1), pharmaceutical manufacturers should be aware that facilities may be more inclined to take advantage of existing syringe dwell times or pursue extended in-syringe stability,[129] which could potentially subject drug products to prolonged contact with unintended materials (e.g., syringe caps, free silicone within syringe barrels, CSTD components).

#### Current and future impact on nursing staff

While nurses reported several benefits of SCOmAbs as anticipated, they also identified challenges that, if addressed during product development, could improve experience with these products and their integration into in-clinic workflows. This was a significant finding in our study, as data from existing literature typically highlights the benefits of reduced administration time and resource requirements without exploring potential burden on nurses in detail. Our study suggests both benefits and drawbacks must be considered, as half of nurses reported that the current manual administration process is burdensome enough to create staffing issues for their facilities and encourage specific strategies to streamline the process. Nurses described that this burden was mostly rooted in the need to be “tethered” to the patient for the duration of each administration, rather than having the freedom to start the administration (i.e., with an infusion pump) and move on to another task, as is typical with IV delivery. This concept is reflected in some studies that show comparatively longer *direct administration times* (i.e., SC injection time versus IV infusion initiation time) for SC vs. IV trastuzumab despite total nurse time being significantly shorter for the SC route.[10,59,79] In these studies, the bulk of nurse time savings with SC delivery was accomplished through elimination or reduction of other IV-related tasks, such as installation/management of venous access, administration of premedications, patient monitoring, and disconnection/disposal of materials. While the degree of tethering varies with IV administration (e.g., patients with existing central venous access do not require catheter insertion, many premedications can be administered orally, etc.), these eliminated IV-related tasks do still require bedside presence, making nurses’ fixation on SC tethering in our study somewhat surprising.

One possible explanation is that SCOmAb delivery provides minimal “hands-free time” compared to IV administration, where the infusion time (sometimes on the order of hours) can offer a break from “tethered time” and is used for multitasking and caring for other patients. For better or worse, multitasking is a substantial element of nursing practice, with studies suggesting that nurses spend an average of 21-29% of their total shift time multitasking, particularly during medication administration.[130,131] Another explanation could be that tethered time for SCOmAb administration requires more ergonomically strained positions compared to other nursing tasks as described above, suggesting that improved injection ergonomics could potentially alleviate this concern.

Beyond the current benefits and limitations of manual delivery, some nurses in our sample noted that further adoption of SCOmAbs could increase strain on infusion center operations and necessitate additional staff and infrastructure to manage it. If such expansion occurred, these nurses felt strongly that their facilities may need to dedicate more staff, training, and facility space specifically to SCOmAb administrations, and potentially utilize syringe pump attachments for administration. Moreover, as additional SC formulations receive regulatory approval, the likelihood increases that patients will receive multiple SCOmAbs as part of combination regimens administered sequentially as separate formulations,[86,132] compounding the per-administration workflow and ergonomic challenges identified in this study across multiple treatments within a single visit. These capacity concerns are echoed by a recent qualitative study of SACT day unit nurses in the UK, who described severe staffing shortages, constrained ability to utilize temporary staff due to specialized training requirements, and increasing patient volumes without corresponding workforce expansion.[133] All things considered, there is no doubt that SC administration of oncology agents offers a variety of tangible benefits compared to IV infusions, and our study results should not be interpreted as refuting the robust body of literature that has clearly established time, resource, and cost savings with SC delivery, and overwhelming patient and HCP preference for this method. Rather, our findings related to pain points with the current process and potential concerns about future expansion should be taken in the context that SCOmAbs *will* continue to be adopted *because* of the benefits they provide, and that improvements should be considered in product development to maximize patient access to these therapies. To realize the full potential of SCOmAbs in terms of time-savings, workflow integration, and patient throughput, especially in light of their increasing prevalence, PFSs and hands-free administration devices might be considered as part of the product presentation and labeled instructions, being mindful of both the benefits and potential complexity these may introduce.

### Potential for home administration

From the study outset, we sought to understand patient and HCP perspectives on home administration of SCOmAbs, including potential enablers and obstacles for home transition. However, we also unexpectedly recruited one patient participant who was currently self-administering a SCOmAb at home and a nurse participant who had extensive experience managing home transitions in her practice. As a result, this section details both the ongoing experiences of the two at-home users in our sample and the projections of current in-clinic users related to at-home treatment.

#### Current home administration practices

The experiences of the patient and nurse in our sample with active home administration programs provided a unique opportunity to corroborate projected benefits and identify practical considerations that may not be apparent from hypothetical discussions alone. Both the patient and nurse in our study confirmed that the primary benefits of home administration were consistent with those projected by the remaining participants in our sample and reported in prior literature, namely time savings, scheduling flexibility, and reduced disruption to daily life. These confirmed benefits align with the findings of Fallowfield et al., who reported that the majority of patients with HER2-positive breast cancer expressed a preference for home delivery of SC trastuzumab, with time savings and convenience cited as the primary motivating factors.[134] In our study, the patient participant further described practical advantages that have not been widely discussed in the context of SCOmAbs, including the ability to obtain her medication and supplies from an outpatient pharmacy and the decoupling of laboratory work from the injection itself, which reduced her total “treatment day” time to approximately 10 minutes. While these specific operational elements may not be generalizable across all SCOmAb regimens, they suggest that home administration has the potential to fundamentally restructure the treatment experience for appropriate patients.

There was similar agreement between our patient and nurse participant with regard to training practices, where both described a comprehensive, one-on-one training process involving nurse demonstration, practice with injection pads, and a supervised self-injection in the clinic before transitioning to the home. This level of individualized training appears to be effective; in a pilot evaluation of a structured self-administration program for SC trastuzumab, Ng et al. reported that 1:1 nurse-led training significantly improved patient confidence in their ability to self-inject, with the vast majority of patients reporting they felt very confident upon completing the program.[135] The nurse participant in our study echoed the importance of a structured approach, noting that candidates for home administration must demonstrate proficiency prior to transitioning, and that follow-up support (e.g., telephone contact after the first at-home injection) was a key element of their program. However, the feasibility of this intensive training model is likely a function of the relatively small number of patients currently performing home administration of SCOmAbs, and potential challenges associated with scaling such programs are considered in the following section.

Although both the patient and nurse in our sample described their existing home administration models as successful, the patient’s account of the self-injection process revealed notable challenges inherent to manual administration. Specifically, she reported using the same needle for both vial draw-up and injection, a practice that can result in needle dulling and increased injection discomfort, and described adopting a “stop and go” technique to manage the sustained syringe plunger pressure required to maintain the recommended injection duration. This latter behavior is particularly noteworthy, as the patient independently developed an unapproved workaround to accommodate the physical demands of the administration process. Such compensatory behaviors are not uncommon among patients who self-administer LVSC therapies, as one study reported that patients self-administering SCIg adopted a variety of self-taught techniques to manage challenges with syringe manipulation, medication preparation, and device operation, many of which deviated from recommended use instructions.[106] Our patient participant’s expressed desire for a self-regulating injection mechanism is also notable, as it aligns with the broader trend that patients who self-administer SC therapies generally prefer automated delivery devices (e.g., AIs) over manual methods, citing ease of use and increased confidence as primary factors.[2] Given that SCOmAb self-administration currently relies on manual syringe injection from vials, a presentation that is less intuitive and more physically demanding than devices specifically designed for patient use, the development of patient-centric delivery devices that simplify the home administration process could meaningfully improve both the feasibility and experience of at-home SCOmAb delivery.

#### Enablers and obstacles for home transition

For those patients in our sample without direct experience with self-administration at home, many still foresaw that doing so instead of their current in-clinic process could offer several benefits, including potential time savings, flexibility, elimination of transportation, and an overall more relaxed environment to receive treatment. Although most research has not specifically evaluated self-administration, our patients’ interest in and perceived advantages of home administration are consistent with prior work, including an examination of hypothetical self-administration of Herceptin Hylecta at home,[134] studies of actual at-home HCP- and self-administered Herceptin Hylecta,[71,136,137], an expanded access study of at-home HCP-administered Phesgo during the COVID-19 pandemic that ultimately led to its approval for home use,[138] evaluations of home treatment services at large cancer institutions,[139–141] and a targeted literature review focused on preferences for in-clinic versus at-home cancer treatment.[142] More recently, the IRAKLIA trial also permitted HCP-administration of SC isatuximab at home via OBDS for patients who had not experienced infusion reactions during prior cycles, representing the first example of device-enabled home administration of a SCOmAb in a clinical trial.[66]

Still, however, patients in our study also expressed hesitations about a transition to self-administration that centered around receiving proper training, troubleshooting administration errors, performing self-administration with a syringe, and abiding by medication storage and sterility requirements. While these concerns were either prospectively mitigated (i.e., through HCP administration) or not raised in the aforementioned studies, they have been reported in the context of self-administration of other therapies outside of oncology. For self-injected biologics used in the treatment of autoimmune conditions, patient training has been reported to be variable and sometimes inadequate, including lack of education on error resolution and troubleshooting steps.[143–145] Although self-administration for these and other conditions is successfully managed by many patients despite suboptimal training, such use cases do not perfectly mirror SCOmAb delivery requirements. More specifically, there are few examples of self-administered SC therapies with dose volumes on the order of SCOmAbs and fewer still that are administered by manual syringe injection. One comparable example is subcutaneous immunoglobulin (SCIg), which is routinely self-administered in similar dose volumes and optionally by “rapid push” via a syringe.[146] While administration of SCIg by manual push may be a viable option for some, several studies have also reported that patients can experience difficulty pushing the syringe plunger due to the viscosity of the product, which could necessitate use of an alternative administration methods.[146–148] Among other issues, concerns about storing medications and supplies, insufficient training, and withdrawing medications from vials to prepare large dose volumes have also been raised among patients receiving SCIg.[106]

By and large, nurses in our sample directly reflected the concerns raised by patients about patient self-administration at home, also citing patient education and training, issue resolution, and the manual syringe administration process itself as potential challenges. As mentioned previously, prior studies on feasibility of home administration actively addressed these concerns by ensuring that home transition was carefully coordinated through a combination of patient initiation, HCP-administration, and/or in-home or remote monitoring,[71,136–141] although one still emphasized the importance of patient education and training on program success.[137] Notably, another specifically described that travel associated with in-home nursing visits was unsustainable at scale, prompting the center to separately evaluate in-home self-administration of SC and intramuscular agents.[141] In the context of self-administration, this constraint may portend that adequate in-home training may be difficult to ensure for all patients, despite all nurses in our study insisting that a thorough training process (either in-clinic or at-home) would be essential for transition to the home. This should be considered in terms of potential adoption of delivery devices to facilitate self-administration at home as well, as lack of scalable patient training would increase the importance of highly intuitive device designs.

Further, some nurses in our study also expressed concerns related to inability to monitor for side effects or adverse reactions if patients are self-administering at home. This is understandable, as active patient monitoring, either through home visits or remotely, has been described as fundamental to Oncology Hospital at Home programs.[149] While the majority of prior studies discussed here utilized in-person monitoring approaches, remote monitoring and the broader use of telehealth has become increasingly common in cancer care, expedited initially by the COVID-19 pandemic.[150–153] Indeed, Dang et al. optionally enrolled a subset of patients in an artificial intelligence-based, remote cardiac surveillance program to monitor for cardiotoxicity associated with at-home Phesgo treatment.[138,154] More broadly, emerging home-based chemotherapy programs in the US have begun to demonstrate that even IV oncology treatments can be safely delivered outside the infusion center with appropriate infrastructure. In a recent pilot, the Mayo Clinic’s Cancer CARE Beyond Walls program successfully delivered 93 home-based IV chemotherapy infusions to 10 patients with no infusion reactions, utilizing a virtual command center, remote patient monitoring, and mobile nursing teams to maintain clinical oversight.[155] While IV home infusion requires substantially more infrastructure than what would be needed for SC manual push administration, this and similar initiatives suggest that the ecosystem required to support home-based oncology care is increasingly feasible, potentially lowering the barrier for SCOmAb home administration as well. Finally, although not specifically mentioned by nurses in our sample, other potential impediments to home administration of cancer treatments have been extensively discussed, most notably administrative and reimbursement barriers in the US.[141,149,156] However, the increasing number of home-based cancer care initiatives that have demonstrated tangible value in recent years have raised credible arguments for reform.[140,157,158] Indeed, in a recent landmark decision, daratumumab and hyaluronidase-fihj became the first oncology injectable approved for patient or caregiver self-administration in Europe, with the labeling update enabling self-administration from the fifth dose for patients with multiple myeloma after appropriate training and healthcare professional assessment.[159] This milestone, which applies to the same manual syringe presentation studied here, underscores the growing regulatory confidence in transitioning SCOmAb administration outside the clinic while also amplifying the importance of the training, patient selection, and device design considerations raised by participants in our study. As developers continue to mature products in this space and as care models also evolve, there is real opportunity to incorporate the feedback and insights from this study, and develop products for home-use SCOmAb administration that can be used safely and effectively.

### Limitations

Our study has several limitations that are worth noting. This was a qualitative study, and while we made every attempt to increase sample diversity, the relatively small sample size still limits overall generalizability of our results. Moreover, although IDI is an effective methodology for understanding current practices and experiences in detail, this approach also has inherent potential for recall and recency bias, as it relies on participant description versus direct observation. Additionally, all coding was performed by a single researcher (MA), which may have introduced individual interpretive bias. While the use of a single experienced coder is common in qualitative studies of this scope, emerging themes and their application were discussed among the broader author team throughout the analysis process to promote consistency and reduce the potential for systematic misinterpretation. The study’s recruiting strategy may have also introduced inconsistencies in responses between stakeholder groups that were difficult to detect. Our recruiting approach was designed to ensure representation from three different stakeholder groups, pharmacists/pharmacy technicians, nurses, and patients, while also maximize the number and geographic diversity of included facilities. As a result, by design, all participants came from different facilities, their reports were treated independently of one another, and their data was then aggregated for each stakeholder group. While this was practical for recruiting efficiency and sample diversity, it also meant that we could not capture different perspectives on the *same workflow* among participants who came from the same facility, which had the potential to produce incongruities. For example, if patients reported that bedside preparation was not performed, we made the reasonable simplifying assumption that preparation was instead performed in pharmacy, although it still is possible that it was indeed performed by nurses in clinical areas but simply out of patient sight (e.g., in a medication room or behind a curtain). An alternative approach could have been to recruit dyads or triads (i.e., pharmacists/technicians, nurses, and patients all from the same facility), but this would have been logistically challenging and yielded results that are potentially less generalizable. Similarly, the requirement for recent experience with SCOmAbs may have selected for more experienced practitioners and potentially underrepresented challenges faced during initial implementation or by those with less familiarity with these agents. This is particularly relevant given the expanding pipeline of SCOmAbs and the likelihood that more facilities and practitioners will encounter these agents for the first time in the near future. Additionally, study data including SCOmAb product names were self-reported by all participants and could not be independently verified against medical records, prescription information, or facility records. However, because all SCOmAbs included in this study rely on similar delivery methods, any reporting inaccuracies are unlikely to have materially affected participant descriptions of their preparation, administration, or injection experiences. On a related note, participant assessments of potential device integration were based on conceptual understanding rather than hands-on experience with specific devices, which may have affected their assessments of fit with current workflows. It is also worth acknowledging that our study captures only a snapshot of practices at a specific time in the evolution of oncology care delivery. The rapid expansion of home-based care models, accelerated by the COVID-19 pandemic and continued through policy changes and technology advancements, means that some facility practices may already be adapting beyond what we observed. Finally, our study included SCOmAbs with dose volumes ranging from 5 to 15 mL, and findings may not apply equally to all SC oncology biologics, particularly those with smaller volumes that may impose fewer physical and workflow constraints. Despite these limitations, to our knowledge, this study represents one of the first systematic examinations of real-world SCOmAb preparation and administration practices across diverse facilities and stakeholder perspectives, providing valuable insights that can inform both current practice optimization and future product development.

## Conclusion

This qualitative study characterizes current practices, challenges, and future considerations for SCOmAb delivery in infusion center settings. Our findings demonstrate that while SCOmAbs offer clear advantages over IV formulations, their integration into clinical practice involves significant workflow considerations that vary by facility size, infrastructure, and staffing resources. The variability observed in preparation practices, administration techniques, and facility-level adaptations highlights both the adaptability of healthcare systems and the need for standardized approaches as SCOmAb utilization continues to expand. Several key insights emerged from this research. While preparation practices are generally well-integrated into pharmacy workflows, inconsistencies in CSTD use and hazardous drug treatment between facilities reflect meaningful variability that manufacturers should anticipate. On the administration side, the ergonomic demands of manual push delivery and the inability of nurses to multitask during injections represent growing concerns as SCOmAb adoption increases, and the creative workarounds already implemented at some facilities, including repurposed syringe pumps, underscore unmet needs in the current administration process. At the same time, patients are naturally intrigued by the idea of self-administration at home, though adequate initiation, easy-to-use delivery devices, and reliable monitoring techniques may be pre-requisites for safe transition. As pharmaceutical manufacturers continue to develop SC formulations of oncology biologics, they should consider how various presentation formats (e.g., PFS, OBDSs, portable infusion pumps, or other delivery devices) might address the identified workflow challenges. Similarly, healthcare facilities should proactively evaluate their infrastructure, staffing models, and training programs to accommodate increasing SCOmAb utilization. Future research should focus on establishing evidence-based best practices for SCOmAb delivery, evaluating the comparative effectiveness of various administration techniques and devices, and further exploring approaches to transition appropriate patients to home administration.

## Acknowledgments

The authors would like to acknowledge Alison Bisch for her contributions to the review of this manuscript. We also thank the study participants for graciously and candidly sharing their practices and personal experiences with the research team.

## Author Contributions

Conceptualization: CF, MA, AR, SB. Data curation: MA. Formal analysis: CF, MA. Funding acquisition: CF, MC, AR, SB. Investigation: MA. Methodology: CF, MA, AR, SB. Project administration: CF, MA, MC. Resources: CF, MC, SB, JW, AR. Supervision: CF, MC, SB. Visualization: CF, MA. Writing - original draft: CF, MA. Writing - review & editing: CF, MA, JW, AR, MC, SB.

## Data Availability

The qualitative data underlying this study consist of interview transcripts containing details about participants and their healthcare facilities. To protect participant privacy as described in the informed consent, these transcripts cannot be shared publicly. De-identified summary data are available from the corresponding author upon reasonable request.

## Supporting Information

S1 Checklist. COREQ (Consolidated Criteria for Reporting Qualitative Research) Checklist for Interviews and Focus Groups

S1 Table. On-Market SCOmAb Injection Attributes

S2 Table. Patient Participants

S3 Table. Nurse and Pharmacy Staff Participants

